# A longitudinal causal graph analysis investigating modifiable risk factors and obesity in a European cohort of children and adolescents

**DOI:** 10.1101/2022.05.18.22275036

**Authors:** Ronja Foraita, Janine Witte, Claudia Börnhorst, Wencke Gwozdz, Valeria Pala, Lauren Lissner, Fabio Lauria, Lucia A Reisch, Dénes Molnár, Stefaan De Henauw, Luis Moreno, Toomas Veidebaum, Michael Tornaritis, Iris Pigeot, Vanessa Didelez

## Abstract

Childhood obesity is a complex disorder that appears to be influenced by an interacting system of many factors. Taking this complexity into account, we aim to investigate the causal structure underlying childhood obesity. Our focus is on identifying potential early, direct or indirect, causes of obesity which may be promising targets for prevention strategies. Using a causal discovery algorithm, we estimate a cohort causal graph (CCG) over the life course from childhood to adolescence. We adapt a popular method, the so-called PC-algorithm, to deal with missing values by multiple imputation, with mixed discrete and continuous variables, and that takes background knowledge such as the time-structure of cohort data into account. The algorithm is then applied to learn the causal relations among 51 variables including obesity, early life factors, diet, lifestyle, insulin resistance, puberty stage and cultural background of 5,112 children from the European IDEFICS/I.Family cohort across three waves (2007-2014). The robustness of the learned causal structure is addressed in a series of alternative and sensitivity analyses; in particular, we use bootstrap resamples to assess the stability of aspects of the learned CCG. Our results suggest some but only indirect possible causal paths from early modifiable risk factors, such as audio-visual media consumption and physical activity, to obesity (measured by age- and sex-adjusted BMI z-scores) six years later.

## Introduction

Childhood obesity is a serious public health problem in many countries [1] leading to severe co-morbidities in later life such as type 2 diabetes, cardiovascular diseases, certain types of cancer, depression and other psychosocial problems [2–4]. Prevention of obesity in children and adolescents seems to be the “only feasible solution” to tackle the obesity epidemic [5]. But prevention strategies need promising targets to achieve any public health effect. However, childhood obesity is a complex disorder that appears to be influenced by an interacting system of individual behaviour, group and societal settings such as family, school or the country-specific infrastructure (e.g. public health system, built environment) [6].

While most investigations focus on single exposure-outcome associations, our approach is to assess the complex interplay of obesity-related factors over the transition from childhood to adolescence by estimating a “cohort causal graph” (CCG), i.e. a causal graph that allows for the longitudinal structure of cohort data, including early life, individual, familial and social aspects using data from the European IDEFICS/I.Family cohort [7]. Our analysis accounts simultaneously for the temporal order of the covariates [8, 9], mixed variable scales and missing values [10]. In addition, we assess the stability and robustness of the estimated causal graph using the bootstrap and further sensitivity analyses. The main aim is to identify plausible causal paths from early modifiable risk factors, such as diet, physical activity (PA), media consumption, subjective well-being and sleep, to body mass index (BMI) six years later. These may suggest or rule out potential targets for future obesity prevention strategies.

## Methods

### Study Population

The IDEFICS/I.Family cohort [7, 11] is a European cohort study initiated with the overall aims to identify and prevent dietary and lifestyle induced health effects in infants, children and adolescents. The baseline survey (B) was conducted in 2007/08 in eight European countries (Belgium, Cyprus, Estonia, Germany, Hungary, Italy, Spain and Sweden) with 16,229 participating children (2 to 9.9 years old). The first follow-up examinations (FU1, conducted in 2009/10) included 13,596 children and applied the same standardised assessments. The second follow-up examinations (FU2, conducted in 2013/14) enrolled 7,105 children who already participated at B or FU1.

### Covariates

We included variables reflecting eating behaviour, lifestyle, social, cultural and environmental factors that are assumed to be related to overweight and obesity across the early life course. A detailed description of all measurements and their units used in our analysis is provided in Table 1 and in the supplement. Some of these variables are time-invariant and would not be targeted by any intervention programme in later childhood, such as region of residence or migration background. Other time-invariant variables might impact a child’s development during pregnancy and as an infant, such as mother’s age at birth or breastfeeding duration; we will refer to these as early life factors. All other variables are time-varying and were measured repeatedly. Age- and sex-specific BMI z-scores (BMI) for children and adolescents were calculated according to the extended IOTF criteria [12]; for simplicity we refer to these as BMI. The homeostatic model assessment (HOMA-IR, short HOMA) index [13] served as a marker for insulin resistance. The diet of the child was measured by a validated FFQ [14] and was classified by an adapted version of the Youth Healthy Eating Index (YHEI) [15]. The YHEI assesses the consumption frequencies of both healthy and unhealthy food as well as eating behaviours, where a higher score indicates a healthier diet [16]. PA was measured by questionnaire, and an audio-visual media consumption score (AVM) was used as proxy for sedentary behaviour. Total sleep duration including nocturnal sleep was estimated based on 24-h dietary recall data at baseline [17] and quantified by self-reports at the two follow-ups. Multiple dimensions of psychosocial well-being were assessed bz questionnaire which was developed for parents’ response on behalf of children and adolescents [18]. Children above the age of 12 completed the questionnaires for themselves. Further details on the study population and used covariates are given in the supplement.

**Table 1:** Variables used in the analysis with units and further explanations. Background knowledge was used to order them into different tiers. Units of continuous variables are given in italics.

| Tiers | Variable/Node | Unit | Comments |
| --- | --- | --- | --- |
| Context | Sex | female, male | Sex of child |
| Context | Region | North, Central, South | Place of residence in one of the following European countries: North (Estonia, Sweden), Central (Belgium, Germany, Hungary), South (Cyprus, Spain) |
| Context | Migrant | no, yes | Children were assumed to have a migrant background if they usually speak with their parents in a language other than the national language of the corresponding country |
| Early life | Mother's age at birth | *years* |  |
| Early life | Total breastfeeding | *months* | Months of breastfeeding, also in combination with other food, prior child's diet was fully integrated into usual household diet |
| Early life | Birthweight | *Gram* |  |
| Early life | Weeks of pregnancy | *weeks* |  |
| Early life | Formula milk | no, yes | Type of feeding prior child's diet was fully integrated into the usual household diet |
| Early life | HH diet | *months* | Month when the child was introduced into the household's diet |
| Early life | Smoking during pregnancy | no, yes | Mother consumed tobacco during pregnancy |
| B, FU1, FU2 | Age | *months* |  |
| B, FU1 | School | kindergarten, school, neither one | Child attended kindergarten/pre-school, school or neither one |
| B, FU1, FU2 | AVM | *h/day* | Audio-visual media consumption score: average hours per day spent with TV, videos, or DVDs, accounting for weekdays and weekends. Hours using the internet per week were only assessed at FU2. |
| B, FU1, FU2 | zBMI | *z-score* | Z-scores of the body mass index [kg/m<sup>2</sup>]. Body weight was measured in fasting state in light underwear on a calibrated scale accurate to 0.1 kg (adapted Tanita BC 420 MA for children ≤6 years, Tanita BC 418 MA for children >6 years, Tanita Europe GmbH, Sindelfingen, Germany); height was measured to the nearest 0.1 cm by a SECA 225 Stadiometer (Seca GmbH & Co. KG., Hamburg, Germany) |
| B, FU1, FU2 | Mother's BMI | *kg/m<sup>2</sup>* | Body mass index of the mother, derived from self-reported weight and height |

| **Tiers** | **Variable/Node** | **Unit** | **Comments** |
| --- | --- | --- | --- |
| B, FU1, FU2 | Daily family meals | no, yes | The family has a meal together at least once a day |
| B, FU1, FU2 | Income | low, middle, high | Country-specific parental income categories, harmonised between countries [56] |
| B, FU1, FU2 | ISCED | low, middle, high | International Standard Classification of Education, highest parental education [57] |
| B, FU1, FU2 | PA | *h/day* | Physical activity measured by questionnaire based on the reported average time spent playing outdoors (hours/week) and the time being in recreation areas or doing sports in a sport club (hours/week) |
| B, FU1, FU2 | Sleep | *h/day* | Nocturnal sleep in hours was assessed by self-reports in FU1 and FU2. The average nocturnal sleep (hours/night) was calculated as the weighted average of reported usual weekday and weekend sleeping times. At baseline, nocturnal sleep was derived based on 24-h dietary recall data where the parents were asked ‘What time did your child go to bed?’ and ‘What time did your child get up?’ |
| B, FU1, FU2 | Well-being | % | Composite sum score; it sums up the answers of 16 items reporting emotional well-being, self-esteem, family relations and peer contacts during the last week, where each item ranges from 0 to 3 points [18, 58] |
| B, FU1, FU2 | YHEI | % | Youth healthy eating score [16] |
| B, FU1, FU2 | HOMA | *z-score* | Z-score of the H<sub>O</sub>meostatic Model Assessment index to quantify insulin resistance; the HOMA-IR index [pg/ml*mg/dl] was calculated from insulin and glucose obtained from blood samples |
| FU2 | Alcohol | no, yes | Ever alcohol drinking in teen's life-time |
| FU2 | Puberty | pre- or early<br>pubertal, pubertal | Pubertal status based on development of voice (boys) and menarche (girls) [59]. Different pubertal stages were displayed in the questionnaire to assist the self-assessment |
| FU2 | Smoking | no, yes | Ever smoking tobacco in teen's life-time |

B: Baseline, FU1: first follow-up, FU2: second follow-up

### Statistical Analysis

For our analyses, only children who participated in all three surveys were considered. Multiple imputation (MI) was applied to avoid loss of study subjects and to reduce potential bias due to missing values [19]; specifically we used tenfold imputation with random forests as implemented in the R-package *mice* [20]. MI assumes that values were missing at random (MAR). To strengthen the plausibility of the MAR assumption, the imputation models were fitted on a larger dataset containing additional variables that contribute to the various scores such as AVM or well-being [19].

To estimate the cohort causal graph (CCG), we applied a method of causal discovery known as PC-algorithm [21]. The algorithm outputs empirically plausible causal directed acyclic graphs (causal DAGs) suggesting direct and indirect causal relations, as shown by directed edges or directed paths. As a DAG represents certain conditional (in)dependencies between variables [22], the PC-algorithm proceeds by investigating conditional independencies in the data using statistical tests, and then determines all DAGs that agree with these independencies. The result is not unique since different DAGs can represent the same conditional independencies, i.e. certain causal structures are indistinguishable. Instead, the algorithm outputs the equivalence class of all DAGs that represent the detected conditional independencies. This class is represented by a so-called completed partially directed acyclic graph (CPDAG) [23] containing directed and undirected edges, where an undirected edge means that both causal directions occur in the equivalence class. The validity of the PC-algorithm relies on the assumptions of causal sufficiency, i.e. absence of latent confounding, and of faithfulness, under which the PC-algorithm consistently selects the true CPDAG [21]. Of note, while the causal interpretation of directed edges or paths in the output of causal discovery algorithms relies on causal sufficiency, which may often be implausible, the absence of such edges and paths can still be interpreted as absence of causal relations even without causal sufficiency.

The PC-algorithm had to be modified for application to multiply imputed cohort data [10, 24, 25]. Further, to account for the cohort structure we used the tiered PC-algorithm *tPC* [26]. This was then combined with functions from *micd* [27] to deal with multiply imputed data containing a mix of categorical and continuous variables. The R packages *micd* and *tPC* are both extensions of *pcalg* [28]. The *tPC*-algorithm outputs a maximally oriented partially directed acyclic graph (MPDAG), which is similar to a CPDAG but can contain more directed edges due to background knowledge [8, 29]. *tPC* determines an MPDAG under the restriction that edges are prohibited from pointing backwards in time which also reduces the number of required statistical tests for conditional independencies. In our analysis we pre-specified the following ordering: region, sex and migration → ISCED and income at baseline → all early life factors → baseline variables → ISCED and income at FU1 → remaining FU1 variables → ISCED and income at FU2 → remaining FU2 variables. Additionally, specific orientations between certain pairs of variables were prohibited, for example from breastfeeding to birth weight. We carried out a number of alternative and sensitivity analyses to check the robustness of the estimated MPDAG against specific analytical choices: (a) while the main analysis used a nominal level of 0.05 for the conditional independence tests, we compared this with a nominal level of 0.1 (MI-0.1); (b) using test-wise deletion (TWD) instead of MI and (c) applying a different, likelihood-based, causal discovery algorithm which uses the EM algorithm for missing values [30]. Moreover, to assess the general stability of the output we drew 100 bootstrap samples from the analysis data, applied to each a single random forest imputation using the same imputation model as in the main analysis, and then estimated 100 bootstrap graphs (BGs). Thus, we can take the frequencies of interesting causal structures in the bootstrap samples as indication of their stability, e.g. specific edges (direct causal links) or indirect links via (partially) directed paths between exposures and outcome. In a directed path, all edges between two nodes are directed, while in a partially directed path, at least one edge between two nodes is undirected. More background on causal graphs and other graph characteristics are described in the supplement.

## Results

### Study Sample

The study sample included 5,112 children who participated in all three surveys. Table 2 shows that children were on average aged 5.9 years at baseline and 11.7 years at FU2. BMI z-scores increased on average by approx. 0.2 standard deviations (SD) over the years (0.32 to 0.55). The overall number of missing values was 15 % with some variables exhibiting very large numbers of missings such as PA at FU2 (50.1 %) (see Figure S1 and Table S1 characteristics after imputation). Diagnostic plots of the multiply imputed data were satisfactory (see Figure S2).

**Table 2:**
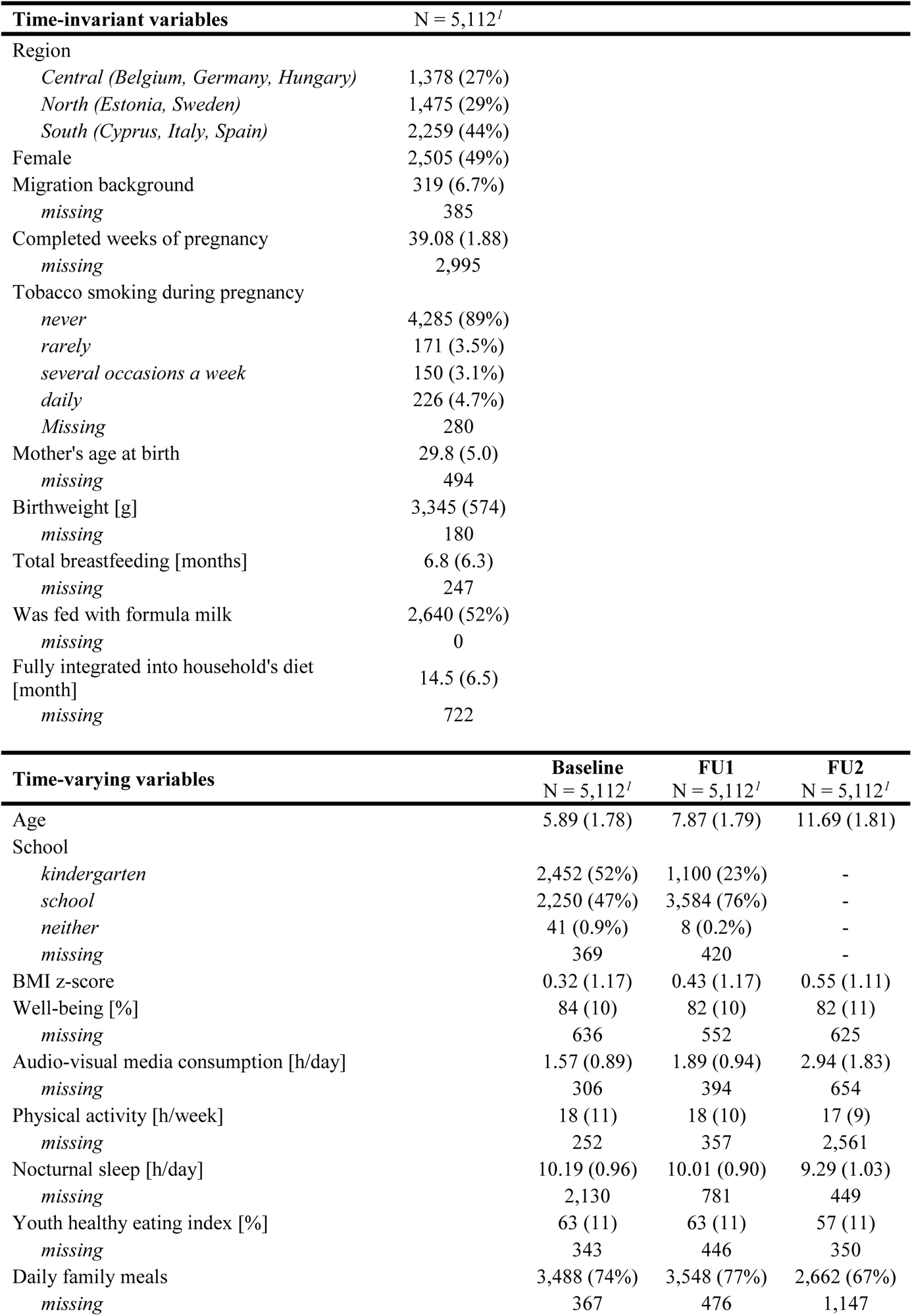
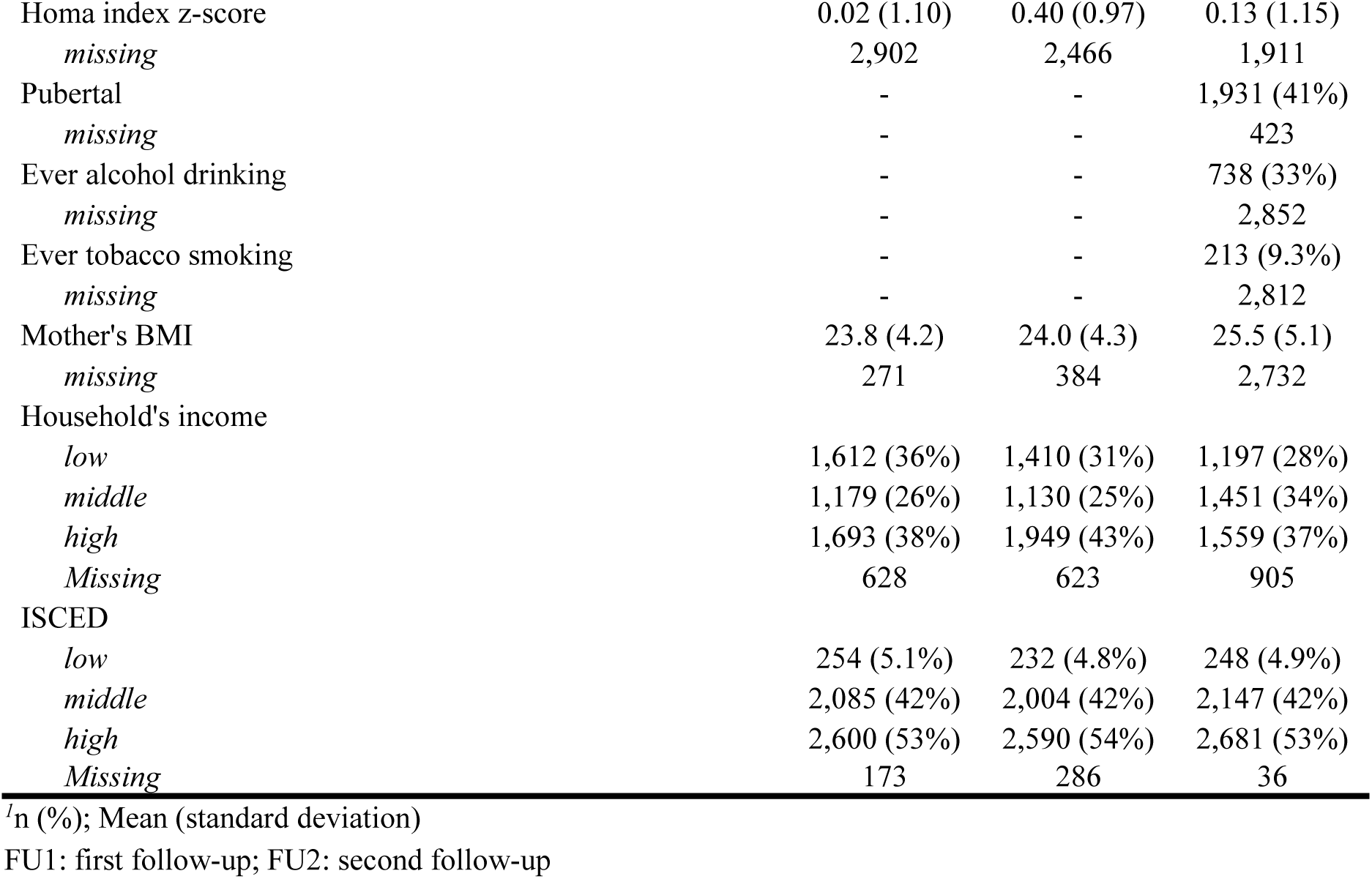
Characteristics of children in the IDEFICS/I.Family cohort participating in all three surveys from 2007 to 2014

| Time-invariant variables |  | N = 5,112<sup>I</sup> |  |  |
| --- | --- | --- | --- | --- |
| Region |  |  |  |  |
|  | *Central (Belgium, Germany, Hungary)* | 1,378 (27%) |  |  |
|  | *North (Estonia, Sweden)* | 1,475 (29%) |  |  |
|  | *South (Cyprus, Italy, Spain)* | 2,259 (44%) |  |  |
| Female |  | 2,505 (49%) |  |  |
| Migration background |  | 319 (6.7%) |  |  |
|  | *missing* | 385 |  |  |
| Completed weeks of pregnancy |  | 39.08 (1.88) |  |  |
|  | *missing* | 2,995 |  |  |
| Tobacco smoking during pregnancy |  |  |  |  |
|  | *never* | 4,285 (89%) |  |  |
|  | *rarely* | 171 (3.5%) |  |  |
|  | *several occasions a week* | 150 (3.1%) |  |  |
|  | *daily* | 226 (4.7%) |  |  |
|  | *Missing* | 280 |  |  |
| Mother's age at birth |  | 29.8 (5.0) |  |  |
|  | *missing* | 494 |  |  |
| Birthweight [g] |  | 3,345 (574) |  |  |
|  | *missing* | 180 |  |  |
| Total breastfeeding [months] |  | 6.8 (6.3) |  |  |
|  | *missing* | 247 |  |  |
| Was fed with formula milk |  | 2,640 (52%) |  |  |
|  | *missing* | 0 |  |  |
| Fully integrated into household's diet [month] |  | 14.5 (6.5) |  |  |
|  | *missing* | 722 |  |  |
| Time-varying variables |  | Baseline<br>N = 5,112<sup>I</sup> | FU1<br>N = 5,112<sup>I</sup> | FU2<br>N = 5,112<sup>I</sup> |
| Age |  | 5.89 (1.78) | 7.87 (1.79) | 11.69 (1.81) |
| School |  |  |  |  |
|  | *kindergarten* | 2,452 (52%) | 1,100 (23%) | - |
|  | *school* | 2,250 (47%) | 3,584 (76%) | - |
|  | *neither* | 41 (0.9%) | 8 (0.2%) | - |
|  | *missing* | 369 | 420 | - |
| BMI z-score |  | 0.32 (1.17) | 0.43 (1.17) | 0.55 (1.11) |
| Well-being [%] |  | 84 (10) | 82 (10) | 82 (11) |
|  | *missing* | 636 | 552 | 625 |
| Audio-visual media consumption [h/day] |  | 1.57 (0.89) | 1.89 (0.94) | 2.94 (1.83) |
|  | *missing* | 306 | 394 | 654 |
| Physical activity [h/week] |  | 18 (11) | 18 (10) | 17 (9) |
|  | *missing* | 252 | 357 | 2,561 |
| Nocturnal sleep [h/day] |  | 10.19 (0.96) | 10.01 (0.90) | 9.29 (1.03) |
|  | *missing* | 2,130 | 781 | 449 |
| Youth healthy eating index [%] |  | 63 (11) | 63 (11) | 57 (11) |
|  | *missing* | 343 | 446 | 350 |
| Daily family meals |  | 3,488 (74%) | 3,548 (77%) | 2,662 (67%) |
|  | *missing* | 367 | 476 | 1,147 |

|  |  |  |  |
| --- | --- | --- | --- |
| Homa index z-score | 0.02 (1.10) | 0.40 (0.97) | 0.13 (1.15) |
| *missing* | 2,902 | 2,466 | 1,911 |
| Pubertal | - | - | 1,931 (41%) |
| *missing* | - | - | 423 |
| Ever alcohol drinking | - | - | 738 (33%) |
| *missing* | - | - | 2,852 |
| Ever tobacco smoking | - | - | 213 (9.3%) |
| *missing* | - | - | 2,812 |
| Mother's BMI | 23.8 (4.2) | 24.0 (4.3) | 25.5 (5.1) |
| *missing* | 271 | 384 | 2,732 |
| Household's income |  |  |  |
| *low* | 1,612 (36%) | 1,410 (31%) | 1,197 (28%) |
| *middle* | 1,179 (26%) | 1,130 (25%) | 1,451 (34%) |
| *high* | 1,693 (38%) | 1,949 (43%) | 1,559 (37%) |
| *Missing* | 628 | 623 | 905 |
| ISCED |  |  |  |
| *low* | 254 (5.1%) | 232 (4.8%) | 248 (4.9%) |
| *middle* | 2,085 (42%) | 2,004 (42%) | 2,147 (42%) |
| *high* | 2,600 (53%) | 2,590 (54%) | 2,681 (53%) |
| *Missing* | 173 | 286 | 36 |

<sup>1</sup>n (%); Mean (standard deviation)
FU1: first follow-up; FU2: second follow-up

### Cohort Causal Graph

The CCG resulting from our main analysis is shown in Figure 1 (see also https://bips-hb.github.io/ccg-childhood-obesity for an interactive graph). Overall the graph was rather sparse with 104 edges linking 51 variables, of which 12 could not be oriented. Focusing on BMI as outcome, there were direct links from region, familial educational level, birthweight and mother’s BMI (B) to BMI (B); in contrast, there were no paths from any of the modifiable risk factors to BMI (B). However, all of these modifiable baseline factors (sleep, AVM, YHEI, PA, well-being) were possible ancestors and hence possible causes of BMI in both follow-ups (cf. Table 3), i.e. they had partially directed paths to BMI. These included paths from all five modifiable baseline risk factors to BMI six years later. For instance, there were five partially directed paths from YHEI (B) to BMI (FU2) (Figure 2). Almost all paths between exposures and BMI (FU2) went through AVM (FU1) and HOMA (FU1, FU2), many also through well-being (FU1) and some through YHEI (B). In the CCG we also see that the exposures themselves were moderately interconnected within the same tier and across time, with many orientations of edges among the exposures at FU1 being undecidable. Note also that most repeated measurements were linked by edges with the notable exception of BMI.

**Figure 1:**
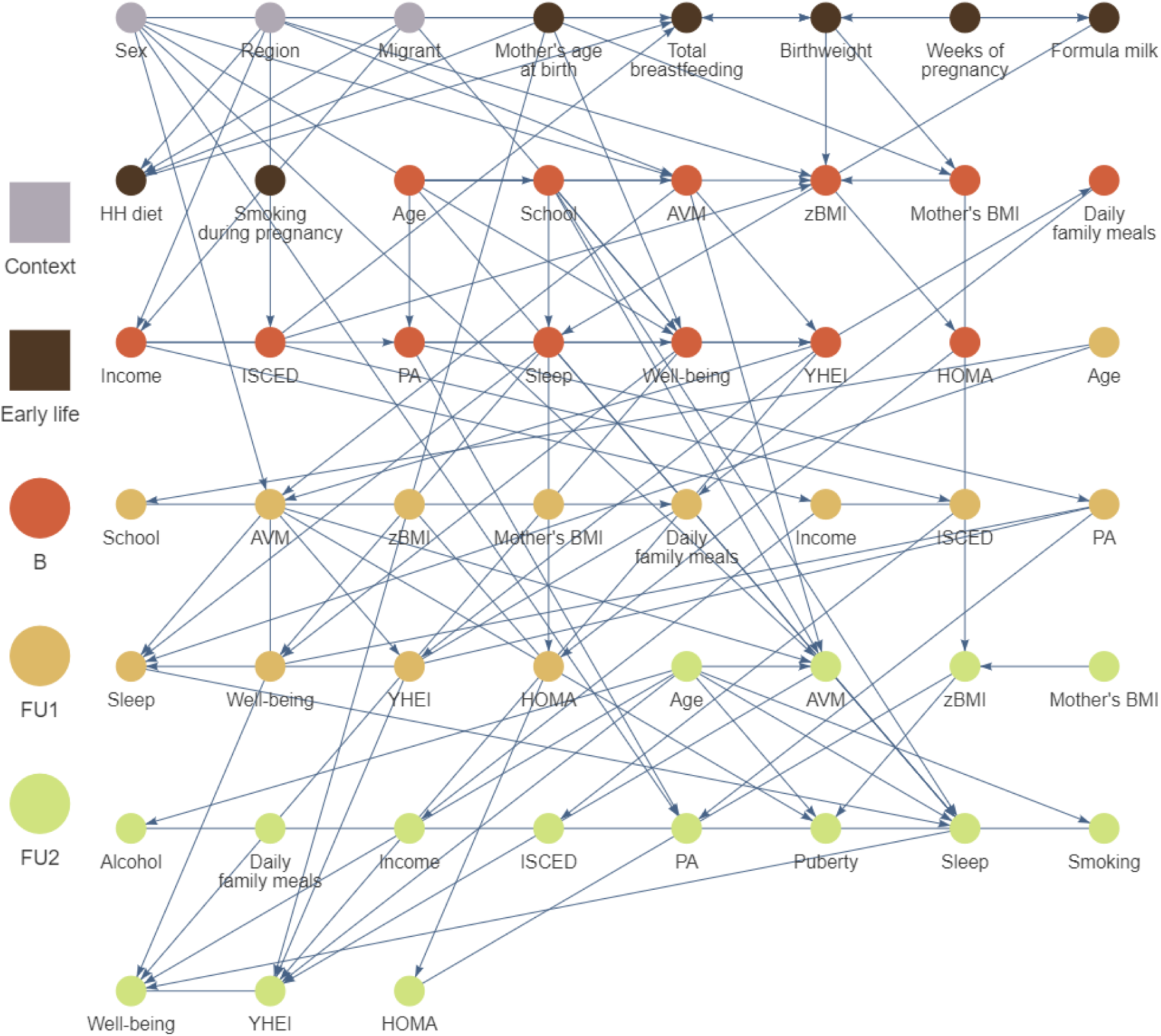
Causal graph of childhood obesity based on N = 5,112 European children and adolescents born between 1997 and 2006 estimated by the tiered PC-algorithm for multiple imputed datasets. The nodes colours correspond to the different stages of the life course. Edges without arrowheads could not be orientated by the algorithm. An overlap of nodes and edges was unavoidable. We advise to look at the interactive graphs here: https://bips-hb.github.io/ccg-childhood-obesity/. AVM: audio-visual media consumption, B: Baseline, FU1: first follow-up, FU2: second follow-up, HH diet: month when the child was introduced into the household’s diet, HOMA: homeostatic model assessment – insulin resistance, ISCED: highest parental education (International Standard Classification of Education), PA: physical activity, YHEI: youth healthy eating index, zBMI: body mass index z-score

**Figure 2:**
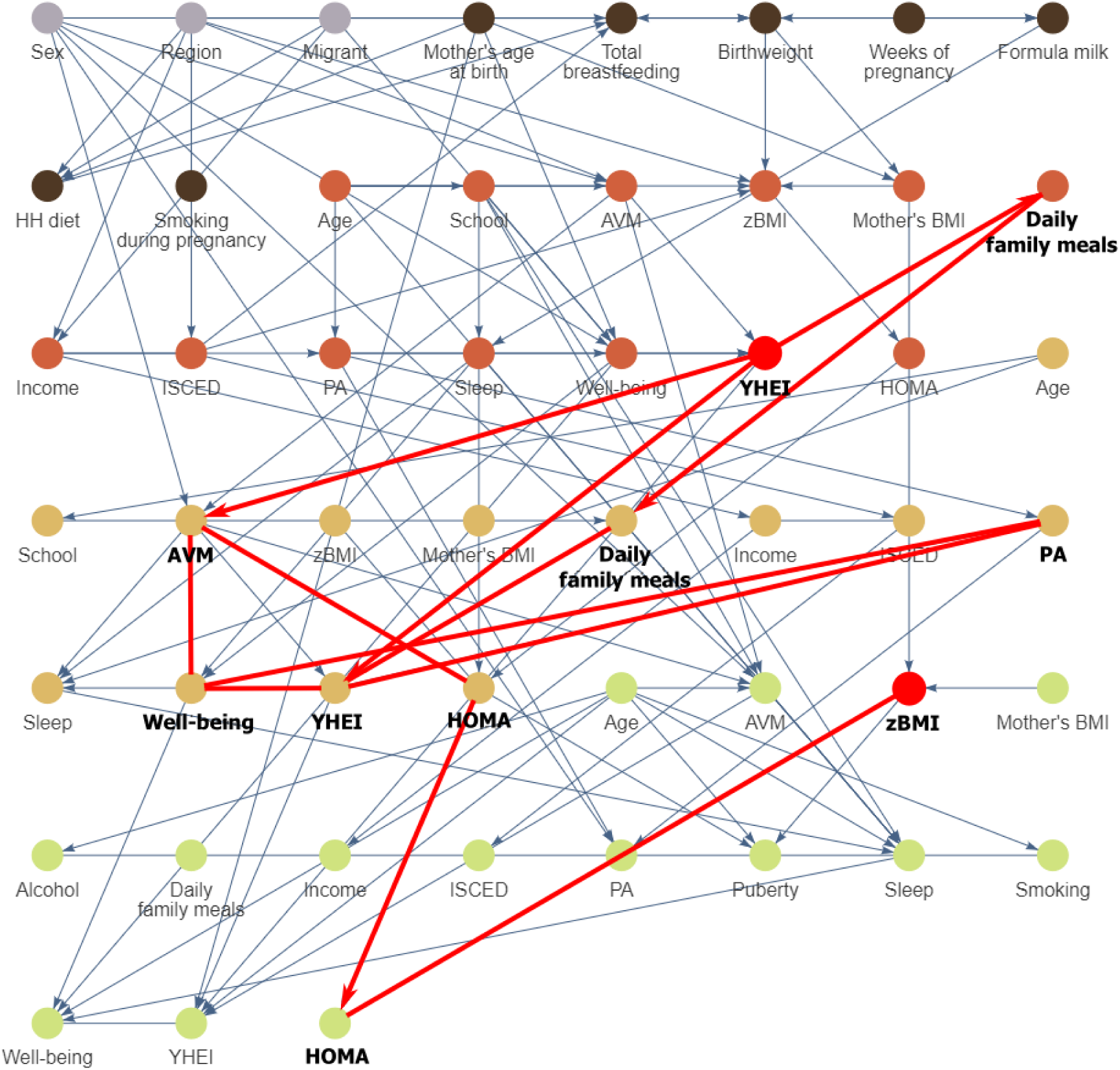
All five possible causal paths between the Youth Healthy Eating Index (YHEI) at baseline and zBMI at the second follow-up (AVM: audio-visual media consumption, PA: physical activity). AVM: audio-visual media consumption, B: Baseline, FU1: first follow-up, FU2: second follow-up, HH diet: month when the child was introduced into the household’s diet, HOMA: homeostatic model assessment – insulin resistance, ISCED: highest parental education (International Standard Classification of Education), PA: physical activity, YHEI: youth healthy eating index, zBMI: body mass index z-score

**Table 3:**
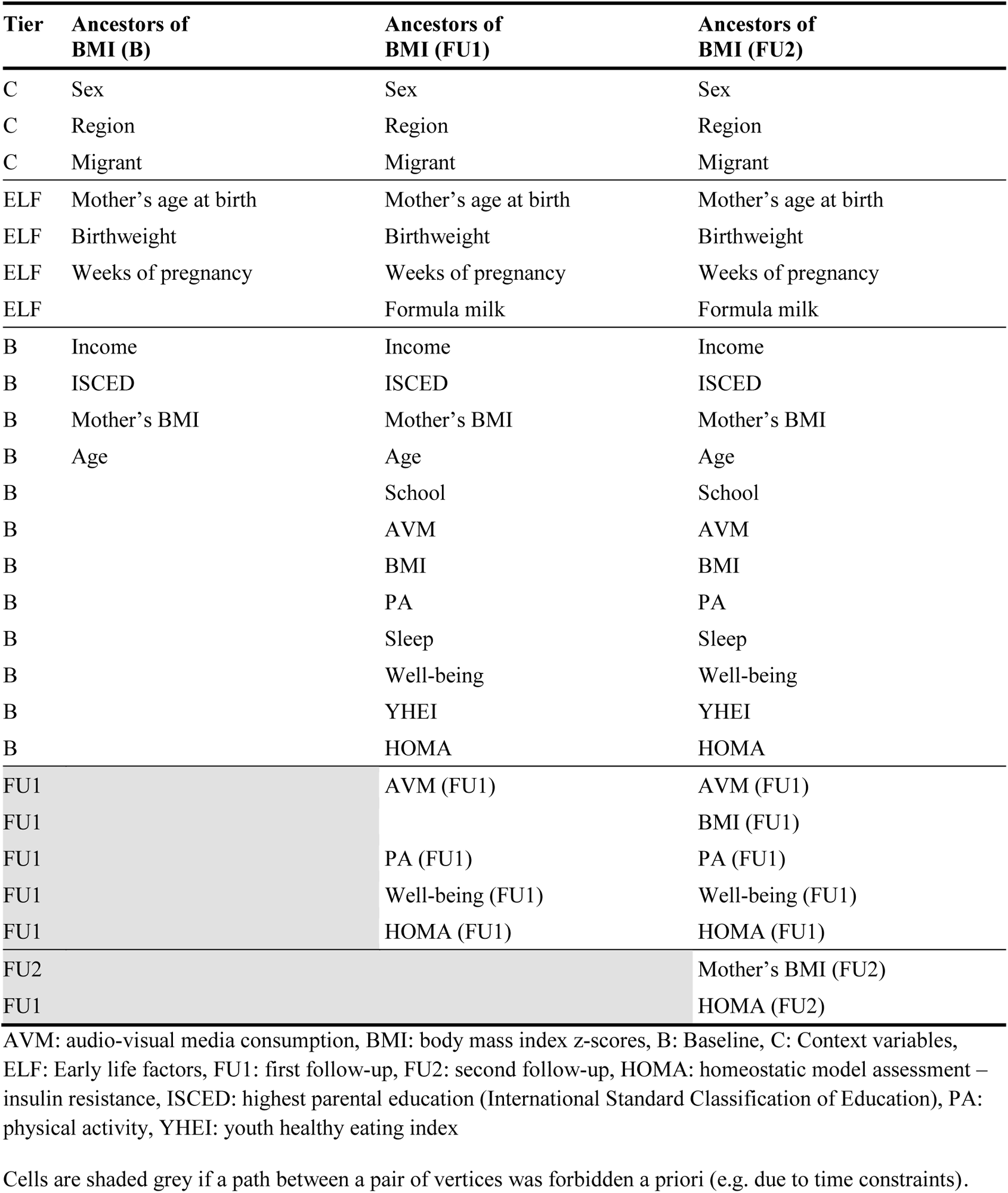
Possible ancestors of BMI at baseline, first and second follow up

### Bootstrap Analysis

We assessed the stability of key features of the main CCG based on 100 BGs. Of the 104 edges in the main CCG, 36 were found to be very stable (in more than 80% of BGs), with a further six edges in more than 70% of BGs (see Table S2). Of these stable edges, 16 were between repeated measures of the same variable, e.g. HOMA.FU1-HOMA.FU2, and 13 emanated from modifiable risk factors. In contrast, 50 edges were clearly unstable, i.e. occurred in 50% or fewer of the BGs. The presence of any paths from exposures to BMI was rather stable. Specifically, we considered directed or partially directed paths from baseline modifiable exposures to later BMI (FU2) (see Table 4). The most frequent were paths from YHEI to BMI (84% of BGs), while paths from sleep duration to BMI were in 75% of the BGs; paths from the other three baseline exposures (well-being, AVM, PA) to BMI occurred in 80% of the BGs. There were usually multiple causal paths found between an exposure and the outcome, and the number of different possible causal paths was high. For instance, the median number of different (partially) directed paths from AVM (B) to BMI (FU2) found in each BG was 20. No BGs ever contained a direct edge from a baseline modifiable exposure to BMI at FU2. Further, we expected the repeated measurements to be linked to each other, which was true for all modifiable risk factors but not for BMI in the main CCG. Table 5 shows patterns between repeated measurements in the main CCG and the BGs. It can be seen for BMI that in 95 BGs the paths B→FU1→FU2 or B→FU1→FU2←B were found despite not being contained in the main CCG.

**Table 4:**
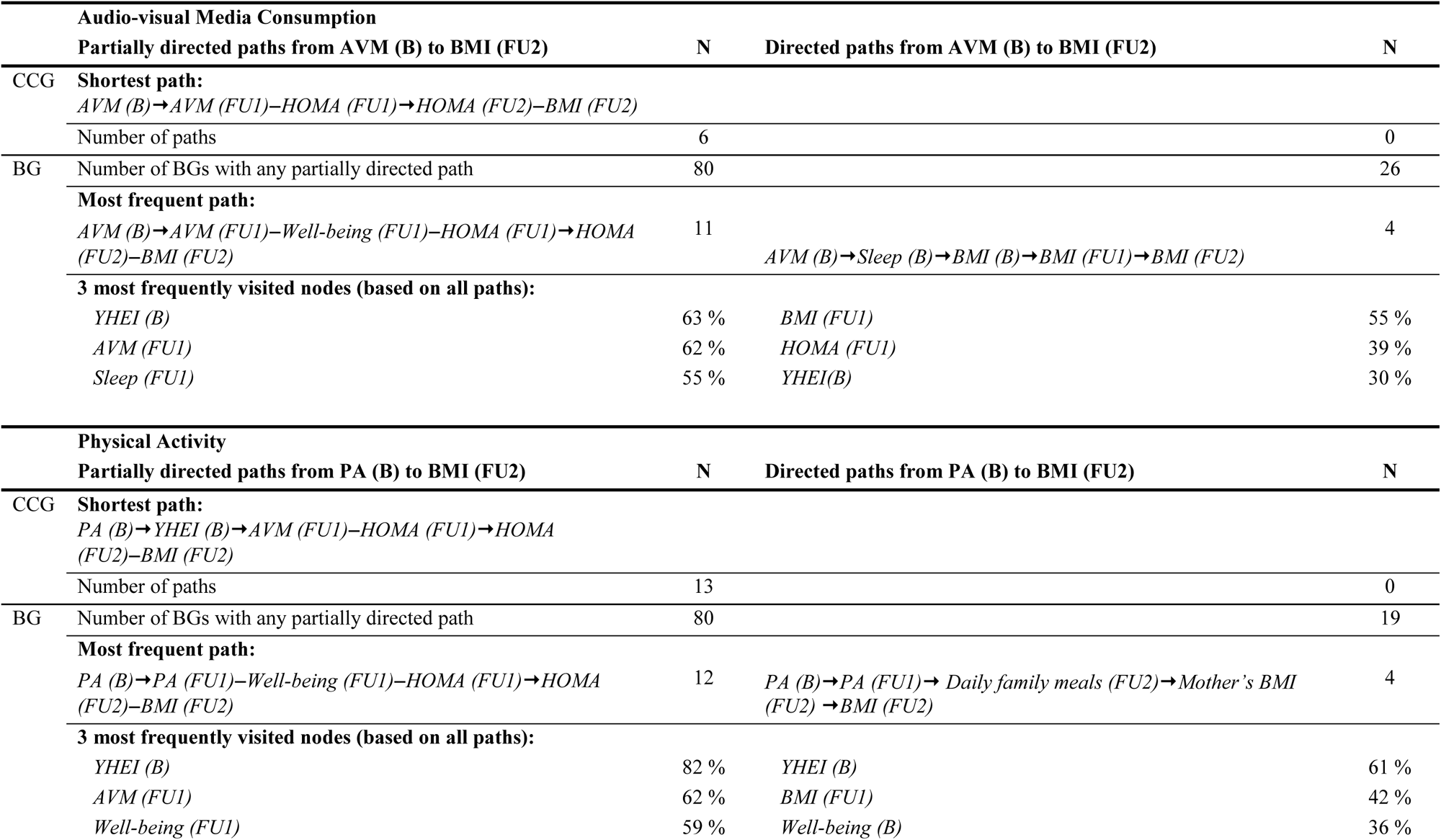
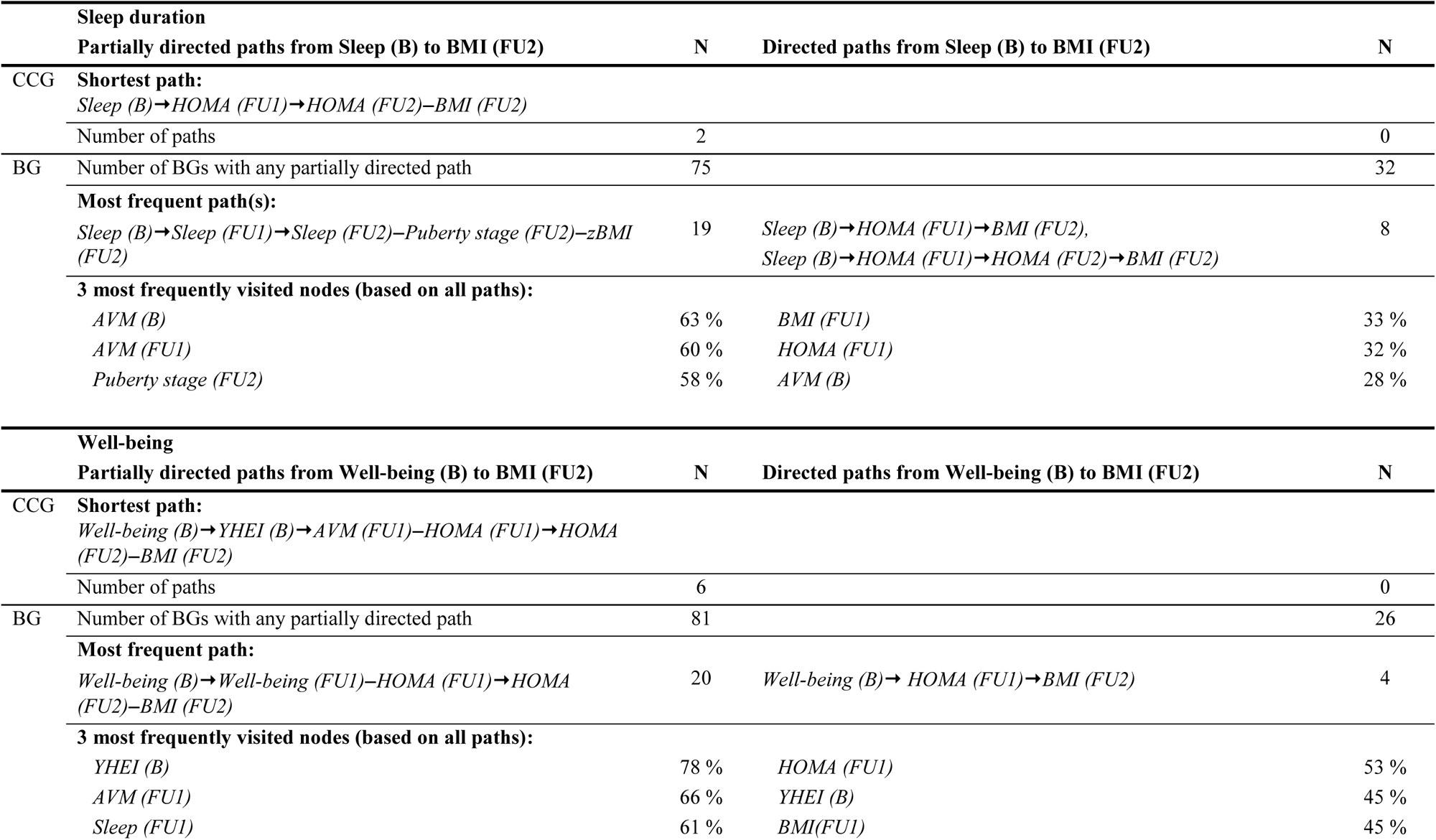
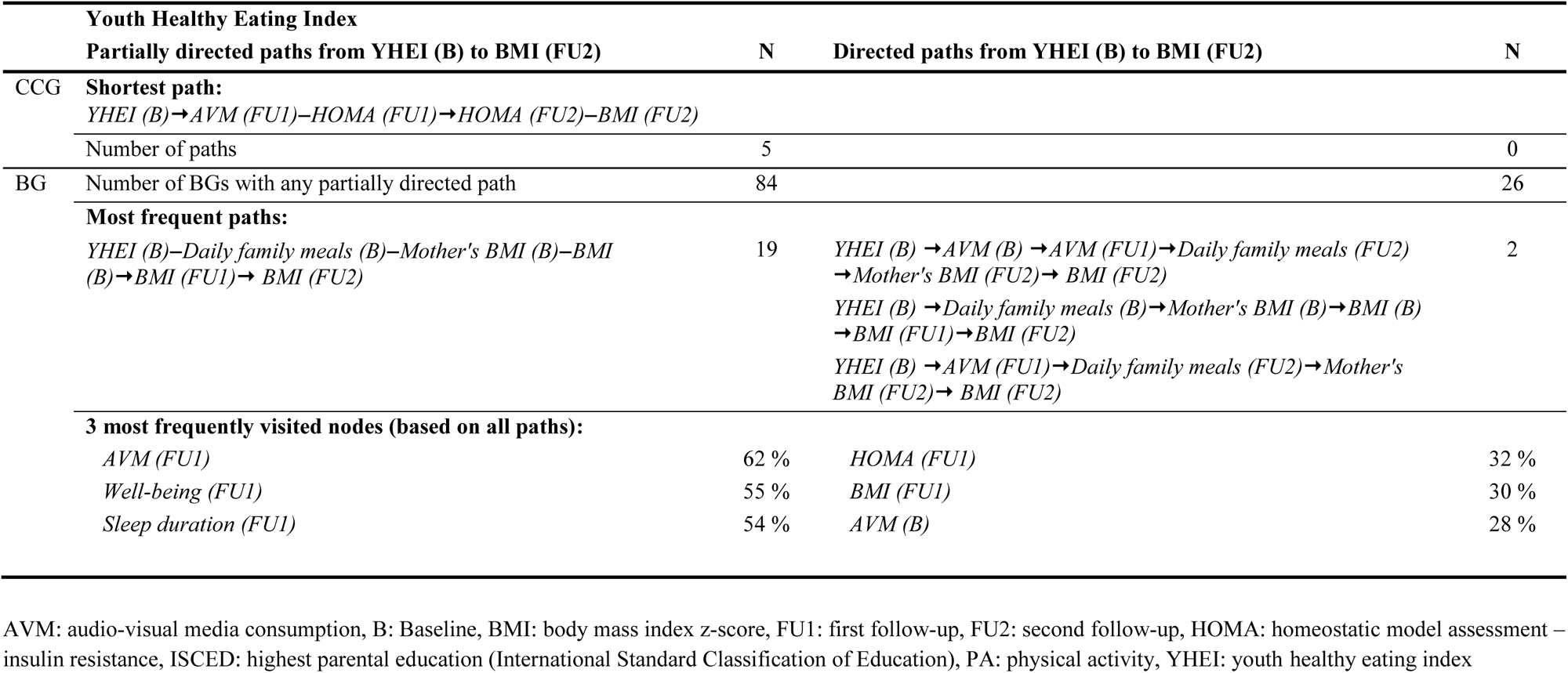
Directed and partially directed paths between modifiable risk factors at baseline and BMI six years later in the original CCG and in 100 Bootstrap graphs (BGs)

**Table 5:** Path patterns between repeated measurements in CCG and Bootstrap graphs.

| Pattern | BMI | AVM | PA | Sleep | Well-being | YHEI | HOMA | Daily family meals | Mother's BMI | Income | ISCED |
| --- | --- | --- | --- | --- | --- | --- | --- | --- | --- | --- | --- |
| None | **2** | - | - | 2 | - | 2 | 0 | 1 | 25 | 0 | 0 |
| B→FU1 | 2 | 2 | 10 | 2 | - | - | 2 | **63** | 0 | 0 | 1 |
| B→FU2 | - | 1 | 2 | - | 1 | 1 | 0 | 2 | **45** | 0 | 0 |
| B→FU1, B→FU2 | - | 1 | 11 | 1 | 2 | 1 | 1 | 34 | 0 | 0 | 0 |
| FU1→FU2 | 1 | 9 | 1 | 13 | 5 | 1 | 0 | 0 | 28 | 35 | 36 |
| B→FU2, FU1→FU2 | - | 1 | 1 | 1 | 1 | - | 0 | 0 | 2 | 5 | 6 |
| B→FU1→FU2 | 82 | 20 | 25 | 37 | 38 | 2 | **59** | 0 | 0 | **57** | **57** |
| B→FU1→FU2←B | 13 | **66** | **50** | **44** | **53** | **93** | 38 | 0 | 0 | 3 | 0 |

AVM: audio-visual media consumption, B: Baseline, BMI: body mass index z-score, FU1: first follow-up, FU2: second follow-up, HOMA: homeostatic model assessment – insulin resistance, ISCED: highest parental education (International Standard Classification of Education), PA: physical activity, YHEI: youth healthy eating index
Bold numbers: path included in main CCG

As suggested by others, BGs tend to be more complex [31, 32], which is also what we found: The BGs contained on average 22 edges more than the CCG in the main analysis. For better comparison with this main CCG, we constructed a graph containing the same number of edges based on the most frequent edges; this resulted in the inclusion of all edges that occurred in more than 44 of BGs (see Figure S6). The (structural) Hamming distance between main CCG and BG44 was 56 (73), indicating that about half of the edges between the two graphs are the same.

### Sensitivity Analyses

Using a larger nominal significance level of 10% (CCG MI-0.1) essentially confirmed the core results from the main graph with only few more edges (Table 6, Figure S3). The CCGs estimated with two alternative methods for missing values (TWD and EM) were with 40 to 50% more edges less sparse than the main graph (cf. Figures S4, S5), where only 20% of the edges in the main analysis were also found in the TWD graph. This was also reflected by the Hamming distances, which was large with 205 for TWD compared to the main CCG. The structural Hamming distance, which additionally counts directional changes, indicated for the MI-0.1 graph that the increase of the nominal level resulted in some undirected edges being directed (e.g., well-being (FU2)→YHEI (FU2)), or vice versa, and others to be re-directed (e.g., the edge between PA (B) and YHEI (B)).

**Table 6:** Characteristics of the discovered graph without singletons

| Characteristics | Main | MI-0.1 | TWD | EM | Avg.BG | BG44 | BG75 |
| --- | --- | --- | --- | --- | --- | --- | --- |
| Number of selected edges | 104 | 113 | 139 | 157 | 126 | 104 | 46 |
| Number of undirected edges | 12 | 13 | 14 | 0 | 12 | 3 | 0 |
| Avg. node degree | 4.8 | 4.9 | 6.0 | 6.2 | 5.4 | 4.2 | 1.8 |
| Max. node degree | 10<sup>1</sup> | 11<sup>2</sup> | 13<sup>3</sup> | 24<sup>4</sup> | 12 | 9<sup>5</sup> | 6<sup>5</sup> |
| Avg. shortest path length | 2.8 | 2.7 | 2.4 | 2.4 | 2.7 | 2.2 | 1.4 |
| Longest shortest path | 9<sup>7</sup> | 8<sup>8</sup> | 7<sup>9</sup> | 7<sup>10</sup> | 8 | 6<sup>11</sup> | 5<sup>12</sup> |
| Hamming distance [60] | - | 19 | 205 | 117 | 88 | 56 | 70 |
| Structural Hamming distance [61] | - | 34 | 214 | 131 | 104 | 73 | 86 |
| Mean edge uncertainty [50] | - | - | - | - | 10.5 | 4.4 | 0.8 |

<sup>1</sup>Region, AVM (FU1), Well-being (B)<sup>2</sup>AVM (FU1), Well-being (B)<sup>3</sup>Migrant<sup>4</sup>Region<sup>5</sup>School (B)<sup>6</sup>Puberty Stage (FU2)<sup>7</sup>Age (FU1) > School (FU1) > Daily family meals (FU1) > YHEI (FU1) > Well-being (FU1) > AVM (FU1) > HOMA (FU1) > HOMA (FU2) > BMI (FU2)<sup>8</sup>Age (FU1) > School (FU1) > Daily family meals (FU1) > YHEI (FU1) > AVM (FU1) > HOMA (FU1) > HOMA (FU2) > BMI (FU2)<sup>9</sup>Weeks of pregnancy (EL) > Daily family meals (B) > Sleep (B) > AVM (B) > Well-being (B) > HOMA (B) > BMI (FU2)<sup>10</sup>Smoking during pregnancy (EL) > Weeks of pregnancy (EL) > Birthweight (EL) > Mother's BMI (B) > AVM (B) > Well-being (B) > Well-being (FU1)<sup>11</sup>Age (B) > School (B) > Well-being (B) > YHEI (B) > PA (B) > PA (FU1)<sup>12</sup>Age (B) > School (B) > Well-being (B) > Well-being (FU1) > Well-being (FU2)
Avg.BG: Occurs on average in each BootG; BG<sub>x</sub>: Summarized bootstrap graph with edges that occurred at least $x$ times in 100 bootstrap replications; EM: structural EM algorithm; Main: multiple imputation with nominal level of 0.05; MI-0.1: multiple imputation with nominal level of 0.1; MEU: Mean edge uncertainty [50]; TWD: test-wise deletion

## Discussion

The estimated CCG suggested rather sparse causal relationships between various variables around childhood obesity, with dependencies of the same measures across time being the strongest and most stable as one might expect. All the individually modifiable risk factors diet, PA, sleep duration, subjective well-being and audio-visual media consumption at baseline were stably found to be possible indirect, but not direct, causes of BMI six years later, mostly via the HOMA index which was closely linked to BMI. Associations between media exposure [33–36], sleep [37–39], PA [37], diet [37], well-being [41] and insulin resistance measured by HOMA were previously found by others and in the IDEFICS/I.Family cohort, partly in smaller subsets and using different variables such as objective accelerometer-based measurements of PA [40–42]. Insulin resistance is strongly associated with obesity, which is reflected by an undirected edge in the CCG. Excess adipose tissue is a known risk factor for insulin resistance; however, normal-weight children may also be affected [43]. From the early life factors, birthweight was a (possible) ancestor of BMI (B, FU1, FU2) and formula milk feeding for BMI (FU1, FU2). High birth weight is known to be associated with childhood obesity [44]; and a recent systematic review describes that there is moderate evidence that breast milk consumption reduces the risk of overweight and obesity at age 2 years and older [45].

Overall, our results suggested that cultural, perinatal and familial variables are potentially more immediate causal influences on obesity, while individually modifiable risk factors play a rather indirect role. Based on the selected CCG, we might hypothesise that early life interventions targeting health behaviours of the child may be insufficient to prevent childhood obesity. This finding is compatible with the view that the causal structure governing childhood health behaviours and outcomes should be considered from a complex adaptive system’s perspective [46–48]. Maitland et al. [49], for example, describe the practical implementation of a “whole of systems” approach.

Using sensitivity analyses we investigated the robustness of the CCG regarding the handling of missing values and used bootstrap samples to assess the stability of learned graph structures. The method for handling missing values is not negligible as more complex and quite different graphs were estimated using TWD or the EM-algorithm instead of MI. Moreover, it was noticeable that the TWD graph, unlike the CCG, was not able to detect edges between repeated measurements. Witte et al. [25] showed that TWD can fail in recovering certain causal structures regardless of the underlying missingness mechanism (MCAR, MAR or MNAR). Further, MI was usually more efficient than TWD, although datasets including variables with mixed measurement scales were more problematic.

We used bootstrap resamples to account for the uncertainty in the selection of the CCG [31, 50, 51]. In interpreting the results it has to be kept in mind that the BGs tended to have more edges than the main CCG, due to spurious dependences induced by sampling with replacement from the given data [31, 32]. We therefore considered the BGs purely as a measure of the stability rather than, say, for estimating edge probabilities. Thus, edge and path frequencies indicate the stability of presence and absence of certain graph structures. While about a third of the learned edges in the main analysis were quite stable, we also found that half of the edges were rather unstable. Similarly, we found that the existence of some paths from early modifiable risk factors to later BMI was quite stable, but the actual paths themselves were very variable, i.e. a particular path may not be selected in more than 20% of BGs. In contrast, the absence of direct links from early modifiable risk factors to later BMI was very stable as these occurred in no BGs. This can be interpreted as the absence of direct causal influences even when the assumption of causal sufficiency is violated.

The main analysis was able to find the expected paths for repeated measurements of HOMA and the modifiable risk factors, but not for BMI, and only partly for daily family meals and mother’s BMI. The BGs runs revealed that missing edges between the repeated measurements of BMI are very rare. The CCG is therefore difficult to explain in this respect. In contrast, the learned CCG suggests the plausible relationship that BMI is conditionally independent of modifiable risk factors given the child’s insulin resistance status (HOMA).

The instabilities that we found through the bootstrap analysis might partly be explained by the rather low sample size for the perhaps rather weak associations, the extra uncertainty due to the high proportion of missing values, and the large intervals between follow-ups. Especially the confidence in specific paths might be rather low which is critical. A greater stability would, for instance, be desirable for subsequent analyses that use a learned causal graph to determine adjustment sets to estimate causal effects [52]. Some graphical rules for identifying adjustment sets just take the adjacent nodes of the exposure into account and others require also the mediators between exposure and outcome, for which reliable knowledge on causal paths is required [53, 54].

Recently, Peterson, Osler & Ekstrom [9] also proposed an extension of the PC-algorithm to include temporal information for inferring a graph from observational data. However, our work is the first application of causal discovery to real-world cohort data accounting jointly for missing values, mixed discrete and continuous variables, and background knowledge such as time-ordering. The required theory and software have only recently been developed [10, 55].

The IDEFICS/I.Family cohort provides a rich source of phenotypes capturing different dimensions of dietary and lifestyle related health aspects repeatedly measured over the early life course. However, a challenge was the choice of variables included in the analysis; these needed to be sufficiently different (i.e. not measuring the same underlying construct) to find meaningful dependencies between the different dimensions of obesity. The further sensitivity analyses (see web page) showed that different choices yielded slightly different selected CCGs, but the overall message remained the same: adolescents’ BMI was not directly affected by earlier behavioural variables, but had indirect, potentially causal, links through AVM (FU1) and HOMA (FU1, FU2).

Further general sources of bias with observational data could also affect our results, such as reporting or selection bias. However, all participating countries adhered to a harmonised protocol and to quality control procedures ensuring high data quality.

## Conclusion

A causal discovery analysis was performed on the IDEFICS/I.Family cohort investigating (causal) dependencies underlying childhood and adolescent obesity in 2 to 16-year-old Europeans. The resulting CCG suggested that cultural, perinatal and familial factors and insulin resistance (HOMA-IR) potentially played a more immediate causal role than individually modifiable risk factors which had stable but only indirect relations with adolescents’ BMI.

## Data Availability

All data produced in the present study are available upon reasonable request to the authors.

## Acknowledgement

This work was done as part of the I.Family Study (http://www.ifamilystudy.eu/) and GrowH! (https://www.growh.eu) and is published on behalf of its consortia. We thank the IDEFICS and I.Family children and their parents for taking the time to participate in this extensive examination programme. We are grateful for the support provided by school boards, headmasters, teachers, school staff and communities, and for the effort of all study nurses and our data managers, especially Claudia Brünings-Kuppe, Sandra Israel-Georgii and Ramona Siebels. We especially thank our colleagues Antje Hebestreit, Maike Wolters, Christoph Buck, Timm Intemann and Heide Busse for their valuable input to realise this interdisciplinary research.

## Supplementary data

Foraita_Supplement.pdf

All CCGs are available as interactive graphs at https://bips-hb.github.io/ccg-childhood-obesity/

## Ethics approval

Ethical approval was obtained from the responsible ethics committees in each country and all research was performed in accordance with the Declaration of Helsinki principles.

## Consent to participate

All children and their parents provided oral and written informed consent, respectively, before examinations and/or the collection of samples, subsequent analysis and storage of personal data and collected samples. Teens older than 12 years were asked to provide their written consent using a simplified version of the consent form. Study subjects and their parents could opt out of each single study component.

