## Supplement for "A longitudinal causal graph analysis investigating modifiable risk factors and obesity in a European cohort of children and adolescents"

### Methods

#### *Study Population*

The IDEFICS/I.Family cohort [1, 2] is a European cohort study initiated with the overall aims to identify and prevent dietary and lifestyle induced health effects in infants, children and adolescents. The baseline survey (B) was conducted from September 2007 to May 2008 in eight European countries (Belgium, Cyprus, Estonia, Germany, Hungary, Italy, Spain and Sweden) with 16,229 participating children (2 to 9.9 years old). Two years (±1 month) later, 13,596 children were included in the first follow-up examinations (FU1, from September 2009 to June 2010). The second follow-up examination (FU2) was conducted from January 2013 to June 2014, in which 7,105 children participated who already participated at B or FU1. The examinations covered a spectrum of parameters following a detailed and standardised study protocol. Parents filled in all questionnaires during B, FU1 and in FU2 if their child was less than 12 years old. Teens aged 12 years or more reported for themselves in FU2. Ethical approval was obtained from the responsible ethics committees in each country. All children and their parents provided oral and written informed consent, respectively, before examinations and/or the collection of samples, subsequent analysis and storage of personal data and collected samples. Teens older than 12 years were asked to provide their written consent using a simplified version of the consent form. Study subjects and their parents could opt out of each single study component. Information on early life factors was obtained from records of routine visits as well as from parental reports. Pregnancy-related questions were posed to biological mothers only.

Information on consumption frequencies was obtained from a food frequency questionnaire (FFQ) with 44 food items (B, FU1) and 59 food items (FU2) from 14 food groups [3]. Daily family meals was further included as binary indicator for healthy food choices and family cohesion [4]. The variables sleep duration and well-being were included as they can indicate a child's stress levels. Nocturnal sleep in hours was assessed by self-reports in FU1 and FU2. The average nocturnal sleep (hours/night) was calculated as the weighted average of reported usual weekday and weekend sleeping times. At baseline, nocturnal sleep was derived based on 24-h dietary recall data where the parents were asked 'What time did your child go to bed?' and 'What time did your child get up?'. The homeostatic model assessment (HOMA-IR, short HOMA) index [5] served as a marker for insulin resistance. Different stages of pubertal status of teens were estimated in FU2 by a self-administered questionnaire based on the development of voice (boys) and menarche (girls) [6, 7]. Unhealthy substance use at FU2 was measured by ever alcohol drinking and ever tobacco smoking. The family's socioeconomic position was assessed by educational level using the International Standard Classification of Education (ISCED)[8] and household income (net income after taxes and deductions), which were harmonised between countries [9]. See Ahrens, Bammann (1), Ahrens, Siani (2) for more details.

*Graph characteristics*

***Adjacent, neighbour, path:*** Two nodes are said to be adjacent, if they are connected by an edge. Such nodes are also referred to as neighbours. A path is an alternating sequence of distinct adjacent nodes and edges as for example $v_0 - v_1 \rightarrow \cdots - v_l$. A directed path proceeds from $v_0$ to $v_l$ along directed edges that point all into the same direction.

***Distance, diameter and average path length:*** The distance between nodes in a graph is commonly defined as the length of the shortest path(s) between these nodes. The distance is infinity when no path exists. The diameter of a graph is defined as the longest path in the graph, which is the maximal distance of any pair of nodes. The average path length of a graph is defined as the average distance between all pairs of nodes. The path length is an indicator for the connectivity of the graph.

***Hamming distance and structural Hamming distance:*** The Hamming distance [10] outputs the minimum number of edge insertions or deletions that are necessary to transform one graph into another graph, where edge directions are not taken into account. Whereas the structural Hamming distance [11] considers edge directions and additionally counts the number of required edge flips that are necessary for a full transformation.

***Root mean squared edge uncertainty (RMSEU):*** The root mean squared edge uncertainty (RMSEU) is a descriptive measure to assess the uncertainty of an undirected graphical model [12]. It reduces the multidimensionality of a graphical model by making use of the edge frequencies of graphs selected from multiple datasets with the same variables. Calculation of this measure was based on 100 bootstrap replications. The boostrap graph BG44 includes edges that were selected in more than 44% of the respective bootstrap graphs. A MEU of 10 (cf. Table 3), for instance, can be obtained for an arbitrary graph where each edge was either selected in 5 % or 95 % of the reruns.

*Background on causal graphs*

***Causal effects:*** A causal effect is defined as the effect of a hypothetical intervention, do($X$=$x$), on an exposure $X$, setting it to $x$ versus $x'$, on the distribution of an outcome $Y$. In the present paper we use the difference in expectation of the outcome as the effect contrast, $E(Y \mid do(X=x)) – E(Y \mid do(X=x'))$. In a causal linear main effects model, $x$ and $x'$ are taken to be one unit apart and the average effect is assumed constant, and equals the slope (no effect modification by included covariates).

***Causal response curve:*** When the exposure is not binary, or when linearity is not appropriate we may want to compare the expected value of a (continuous) outcome $Y$ across different interventional values of $X$, i.e. we want to estimate $E(Y \mid do(X=x))$ as a function of the continuous or multi-valued $X$. We denote the function $E(Y \mid do(X=x))$ as causal response curve (aka ‘expected outcome under hypothetical interventions (EOHI)’).

***Key structural assumptions:*** The estimation of causal effects or causal response curves typically relies on the following key assumptions: (1) causal consistency, meaning that an intervention on the exposure must be well defined such that what we actually observed would have been observed if the exposure value had be set to its value by the intervention; (2) there is no unobserved confounding so that the covariates included in the analysis are sufficient to adjust for any confounding; (3) positivity, meaning that each individual could have in principle be subject to any other exposure value (within the range being compared) by a corresponding intervention.

***Causal DAG:*** A causal directed acyclic graph (causal DAG) consists of nodes representing the variables and directed edges representing direct causal relations; it has no cycles. More precisely, it is the absence of edges that imply the absence of direct causal effects, and consequently the absence of any directed path from $X$ to $Y$ implies the absence of a total (overall) causal effect of $X$ on $Y$. Probabilistically, a causal DAG implies conditional independencies between variables (the causal Markov properties) which can be read off using d-separation [13].

***CPDAG:*** A completed partially directed acyclic graph represents the equivalence class of DAGs, i.e. the set of DAGs that encode the same conditional independencies but not necessarily the same causal relations. For instance $X \rightarrow Y \rightarrow$ Z implies the same conditional independence as $X \leftarrow Y \leftarrow$ Z, even though the causal meaning is very different; the corresponding CPDAG is $X$ – $Y$ – Z. An undirected edge in a CPDAG means that the equivalence class contains at least one DAGs where the edge is directed in one direction and at least one other DAGs where it is directed in the reverse direction. Causal discovery methods that solely rely on conditional independencies found in the data cannot distinguish between different DAGs contained in a CPDAG, i.e. without any external information (or randomization or parametric assumptions) we cannot distinguish $X \rightarrow Y \rightarrow$ Z from $X \leftarrow Y \leftarrow$ Z; thus the only information that the CPDAG $X$ – $Y$ – Z carries is that there is no direct causal relation between $X$ and $Z$ and that $X \rightarrow Y \leftarrow$ Z can be excluded. The DAGs contained in a CPDAG can be obtained by finding all possible edge orientations for the undirected edges such that no cycles and no new V-structures ($X \rightarrow Y \leftarrow$ Z) are created. Asymptotically, the PC-algorithm outputs a CPDAG, but for finite-samples this cannot be guaranteed.

***MPDAG:*** A maximally oriented partially directed acyclic graph is a subset of an equivalence class of DAGs, i.e. a subset of the DAGs contained by a CPDAG; the subset is obtained by adding background knowledge on absence of presence of edges to the conditional independencies. For example, if we knew that $X$ is in time before $Z$ (in addition to $X$ and $Z$ being conditionally independent given $Y$) then we obtain that either $X \leftarrow Y \rightarrow$ Z or $X \rightarrow Y \rightarrow$ Z must hold, which are summarized in the MPDAG $X$ – $Y \rightarrow$ Z.

***Faithfulness:*** The assumption that every conditional independence in the data corresponds to the absence of some edge (and thus to some d-separation) in the underlying causal DAG is known as faithfulness. It can be violated if, for instance, a positive and a negative effect along different pathways cancel out each other exactly.

***Causal sufficiency:*** The assumption that the observed variables can be represented in a causal DAG without additional latent variables (nodes) being common causes of two or more observed nodes is known as causal sufficiency. This is a strong assumption, but while approaches exist to relax causal sufficiency, these are more time-consuming, much more difficult to interpret and have not yet been generalized for time-ordered data. The output of the PC-algorithm can still be interpreted in terms of conditional independencies even in the absence of causal sufficiency. The absence of edges can then still be interpreted as the absence of direct causal relations (under the assumption of faithfulness).

***PC-algorithm:*** The PC-algorithm is named after **P**eter Spirtes and **C**lark Gylmour; it proceeds by determining conditional independencies in the data and then finding a CPDAG that is compatible with the independencies. With perfect conditional independence information, the PC-algorithm is valid (sound and complete) under the assumptions of faithfulness and causal sufficiency. Under additional assumptions on the underlying data generating mechanism it is also consistent [14-16]

***tPC:*** The tiered PC-algorithm [17, 18] is a variant of the PC-algorithm and uses additional prior-knowledge on a partial tiered ordering of the variables (nodes) to exclude certain edge directions. Its output is an MPDAG (which cannot be guaranteed for finite samples). A recent tutorial [19] describes how to apply tPC for causal discovery on cohort data with missing data.

***MICD:*** Multiple imputation causal discovery is described in [18, 20] and provided as R-package [21]. It proceeds by first creating multiply imputed data sets. Conditional independence tests are then performed separately on and then pooled across the multiple datasets. The resulting test decisions are entered into the (t)PC-algorithm resulting in an MPDAG.

***Multiset of causal effects:*** An equivalence class of DAGs or MPDAG contains possibly many different causal DAGs. For example, $X - Z \rightarrow Y$ contains (i) $X \rightarrow Z \rightarrow Y$ and (ii) $X \leftarrow Z \rightarrow Y$; in (i) the effect of $X$ on $Y$ could be non-zero, in (ii) it would be zero. Also, in (i) we would not adjust for $Z$ when estimating the effect of $X$ on $Y$ as it is a mediator, while in (ii) we should adjust for $Z$ as it is a confounder of the $X - Y$ relation (this would only be relevant if there are further paths from $X$ to $Y$ in the graph). This example shows that based on an MPDAG we may find different causal effects and different adjustment sets for the same exposure-outcome pair as we need to allow for the different causal DAGs that cannot be distinguished. Thus, instead of estimating a single causal effect, we estimate a multiset of causal effects, one value for each DAG contained in the MPDAG.

### Results

**Table S1:** Characteristics of children in the IDEFICS/I.Family cohort participating in all three surveys from 2007 to 2014 in 10 datasets imputed by chained equations using random forest

| Time-invariant variables | N = 51,120[1] |
|---|---|
| Region | |
| *Central (Belgium, Germany, Hungary)* | 27 % |
| *North (Estonia, Sweden)* | 29 % |
| *South (Cyprus, Italy, Spain)* | 44 % |
| Female | 49 % |
| Migration background | 6.5 % |
| Completed weeks of pregnancy | 39.24 (1.60) |
| Tobacco smoking during pregnancy | |
| *never* | 89 % |
| *rarely* | 3.4 % |
| *several occasions a week* | 3.0 % |
| *daily* | 4.5 % |
| Mother's age at birth | 29.8 (4.9) |
| Birthweight [g] | 3,344 (571) |
| Total breastfeeding [months] | 6.7 (6.3) |
| Was fed with formula milk | 52 % |
| Fully integrated into household's diet [month] | 14.4 (6.3) |

*To be continued on the next page.*

| **Time-varying variables** | **Baseline** N = 51,120[1] | **FU1** N = 51,120[1] | **FU2** N = 51,120[1] |
|---|---|---|---|
| Age | 5.89 (1.78) | 7.87 (1.79) | 11.69 (1.81) |
| School | | | |
| *kindergarten* | 52 % | 23 % | - |
| *school* | 47 % | 77 % | - |
| *neither* | 0.8 % | 0.2 % | - |
| BMI z-score | 0.32 (1.17) | 0.43 (1.17) | 0.55 (1.11) |
| Well-being [%] | 84 (9) | 82 (10) | 82 (10) |
| Audiovisual media consumption [h/day] | 1.57 (0.87) | 1.89 (0.92) | 2.93 (1.78) |
| Physical activity [h/week] | 18 (10) | 18 (10) | 17 (9) |
| Nocturnal sleep [h/day] | 10.20 (0.89) | 10.00 (0.86) | 9.28 (1.00) |
| Youth healthy eating index [%] | 63 (11) | 63 (11) | 57 (10) |
| Daily family meals | 75 % | 78 % | 69 % |
| Homa index z-score | 0.05 (1.07) | 0.42 (0.94) | 0.09 (1.19) |
| Pubertal | - | - | 41 % |
| Ever alcohol drinking | - | - | 26 % |
| Ever tobacco smoking | - | - | 6.2 % |
| Mother's BMI | 23.7 (4.2) | 24.0 (4.2) | 24.9 (4.9) |
| Household's income | | | |
| *low* | 37 % | 31 % | 28 % |
| *middle* | 26 % | 25 % | 36 % |
| *high* | 37 % | 43 % | 36 % |
| ISCED | | | |
| *low* | 5.0 % | 4.6 % | 4.9 % |
| *middle* | 42 % | 42 % | 42 % |
| *high* | 53 % | 54 % | 53 % |

[1] %; Mean (SD)

**Table S2:** Selected edges in the main graph and their selection frequency in the bootstrap graphs.

| Edges | | | Edges | | |
|---|---|---|---|---|---|
| from | to | % | from | to | % |
| Age (B) | School (B) | 100 | Weeks of pregnancy | Birthweight | 56,5 |
| Age (FU2) | Puberty (FU2) | 100 | Mother's age at birth | Mother's BMI (B) | 56 |
| HOMA (B) | HOMA (FU1) | 100 | HOMA (FU1) | Puberty (FU2) | 55 |
| Income (FU1) | Income (FU2) | 100 | YHEI (B) | AVM (FU1) | 54 |
| Migrant | Income | 100 | Mother's age at birth | HH diet | 53 |
| Region | Income | 100 | Well-being (B) | YHEI (B) | 53 |
| Region | ISCED | 100 | Age (FU2) | YHEI (FU2) | 52 |
| Age (FU1) | School (FU1) | 99 | BMI (FU2) | HOMA (FU2) | 52 |
| Age (FU2) | Alcohol (FU2) | 99 | PA (FU1) | Well-being (FU1) | 51 |
| ISCED (FU1) | ISCED (FU2) | 99 | Region | Birthweight | 50 |
| Sex | AVM (FU1) | 98 | Daily family meals (FU1) | YHEI (FU1) | 49 |
| Daily family meals (B) | Daily family meals (FU1) | 97 | Income | PA (B) | 49 |
| HOMA (FU1) | HOMA (FU2) | 97 | YHEI (FU1) | Daily family meals (FU1) | 49 |
| Well-being (FU1) | Well-being (FU2) | 97 | HOMA (FU2) | BMI (FU2) | 48 |
| AVM (FU1) | AVM (FU2) | 96 | Mother's BMI (B) | BMI (FU2) | 47 |
| PA (B) | PA (FU1) | 96 | YHEI (B) | Daily family meals (B) | 46,5 |
| YHEI (B) | YHEI (FU1) | 96 | Sleep (B) | Sleep (FU2) | 46 |
| YHEI (FU1) | YHEI (FU2) | 96 | BMI (B) | HOMA (B) | 45 |
| School (B) | AVM (FU2) | 95 | YHEI (FU1) | Well-being (FU1) | 44 |
| Sleep (FU1) | Sleep (FU2) | 95 | HOMA (FU1) | AVM (FU1) | 42,5 |
| YHEI (B) | YHEI (FU2) | 95 | AVM (FU2) | YHEI (FU2) | 41,5 |
| Mother's BMI (B) | BMI (B) | 94 | PA (FU1) | YHEI (FU1) | 40 |
| Sex | AVM (FU2) | 94 | PA (B) | YHEI (B) | 39 |
| School (B) | Sleep (FU2) | 93 | AVM (FU2) | Sleep (FU2) | 37,5 |
| Well-being (B) | Well-being (FU1) | 93 | Age (B) | PA (B) | 37 |
| Age (FU2) | Sleep (FU2) | 92 | AVM (B) | YHEI (B) | 37 |
| School (FU1) | Daily family meals (FU1) | 91 | Well-being (FU1) | PA (FU1) | 37 |
| AVM (B) | AVM (FU1) | 89 | Sex | Well-being (B) | 36 |
| Age (B) | AVM (B) | 88 | Smoking (FU2) | Alcohol (FU2) | 36 |
| Sex | AVM (B) | 88 | AVM (FU1) | Well-being (FU1) | 34,5 |
| YHEI (B) | YHEI (FU2) | 95 | Income | ISCED | 34 |
| Sex | PA (FU2) | 86 | Age (FU2) | Well-being (FU2) | 31 |
| Birthweight | BMI (B) | 85 | Migrant | Well-being (B) | 30 |
| School (B) | Sleep (B) | 85 | Age (B) | BMI (B) | 27 |
| Sleep (B) | Sleep (FU1) | 84 | Well-being (FU1) | YHEI (FU1) | 27 |
| Age (FU2) | Smoking (FU2) | 82 | AVM (FU1) | YHEI (FU1) | 26,5 |
| School (B) | Well-being (B) | 82 | Formula milk | Sleep (B) | 26 |
| Weeks of pregnancy | Formula milk | 79 | HOMA (FU1) | BMI (FU1) | 25 |
| Mother's BMI (FU2) | BMI (FU2) | 78 | Mother's age at birth | Well-being (B) | 24 |
| PA (FU1) | PA (FU2) | 77 | Region | AVM (B) | 24 |
| Region | Total breastfeeding | 77 | Well-being (FU1) | Sleep (FU1) | 22,5 |
| BMI (FU1) | HOMA (FU1) | 74 | ISCED | Total breastfeeding | 22 |
| Age (FU2) | AVM (FU2) | 70 | Region | BMI (B) | 22 |
| AVM (B) | AVM (FU2) | 69 | Sleep (B) | HOMA (FU1) | 21 |
| Sex | Birthweight | 69 | Income (FU1) | ISCED (FU1) | 20,5 |
| Formula milk | Total breastfeeding | 67,5 | Migrant | HH diet | 20 |
| ISCED | Income | 66 | ISCED (FU1) | Income (FU1) | 19,5 |
| PA (B) | PA (FU2) | 64 | AVM (FU1) | Sleep (FU1) | 18 |
| Region | Formula milk | 64 | PA (B) | Well-being (B) | 17 |
| Age (FU1) | Sleep (FU1) | 63 | Well-being (FU1) | AVM (FU1) | 15,5 |
| Alcohol (FU2) | Smoking (FU2) | 63 | Well-being (FU2) | YHEI (FU2) | 13,5 |
| BMI (FU2) | Puberty (FU2) | 61,5 | YHEI (FU1) | PA (FU1) | 13 |
| Birthweight | Mother's BMI (B) | 60 | Sleep (B) | Well-being (FU1) | 10 |
| Income | Income (FU1) | 60 | AVM (FU1) | HOMA (FU1) | 9,5 |
| Region | HH diet | 60 | Mother's age at birth | YHEI (FU2) | 9 |
| HH diet | Total breastfeeding | 59 | Sleep (FU2) | Well-being (FU2) | 8,5 |
| ISCED | ISCED (FU1) | 58 | ISCED | BMI (B) | 8 |
| Well-being (B) | Well-being (FU2) | 57 | Age (B) | Sleep (FU2) | 7 |
| Weeks of pregnancy | Birthweight | 56,5 | YHEI (FU2) | Well-being (FU2) | 3,5 |

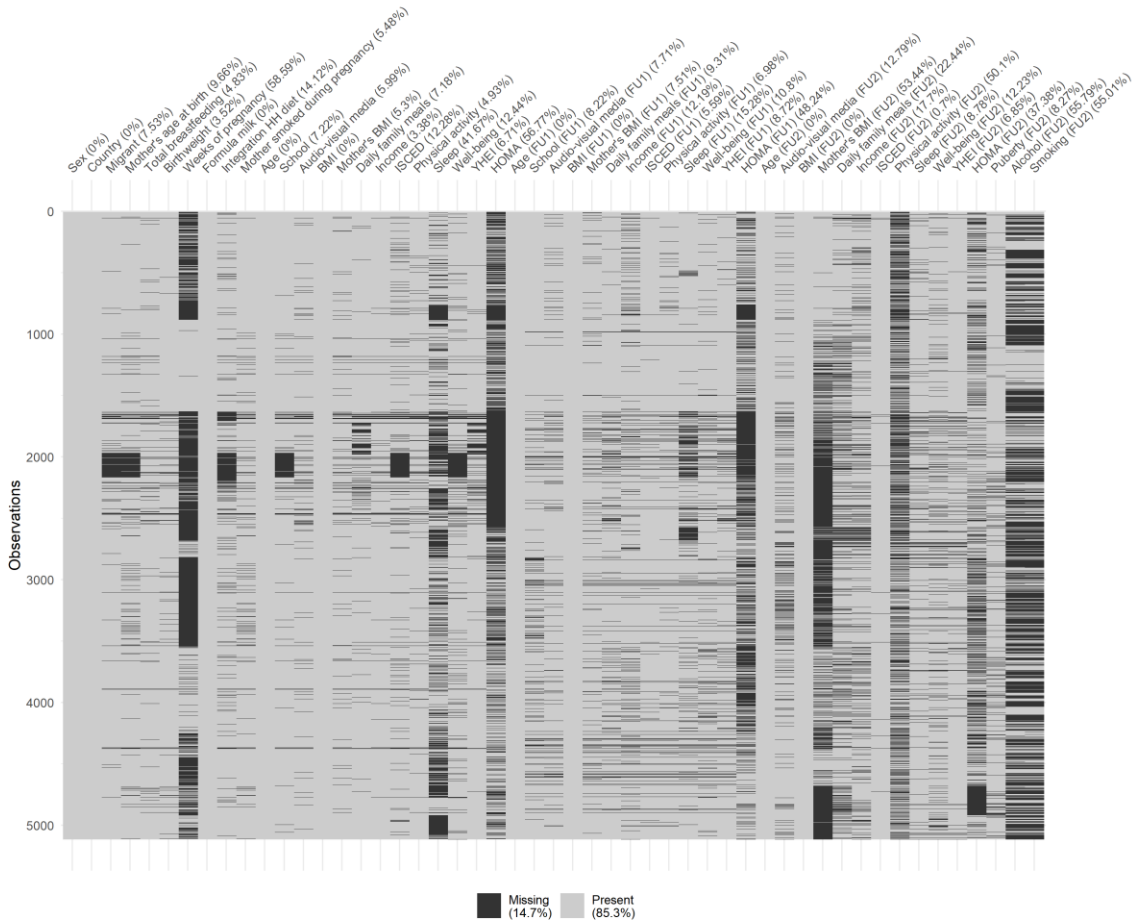


**Figure S1:** Missing values in the cohort dataset where black cells indicate a missing observation.

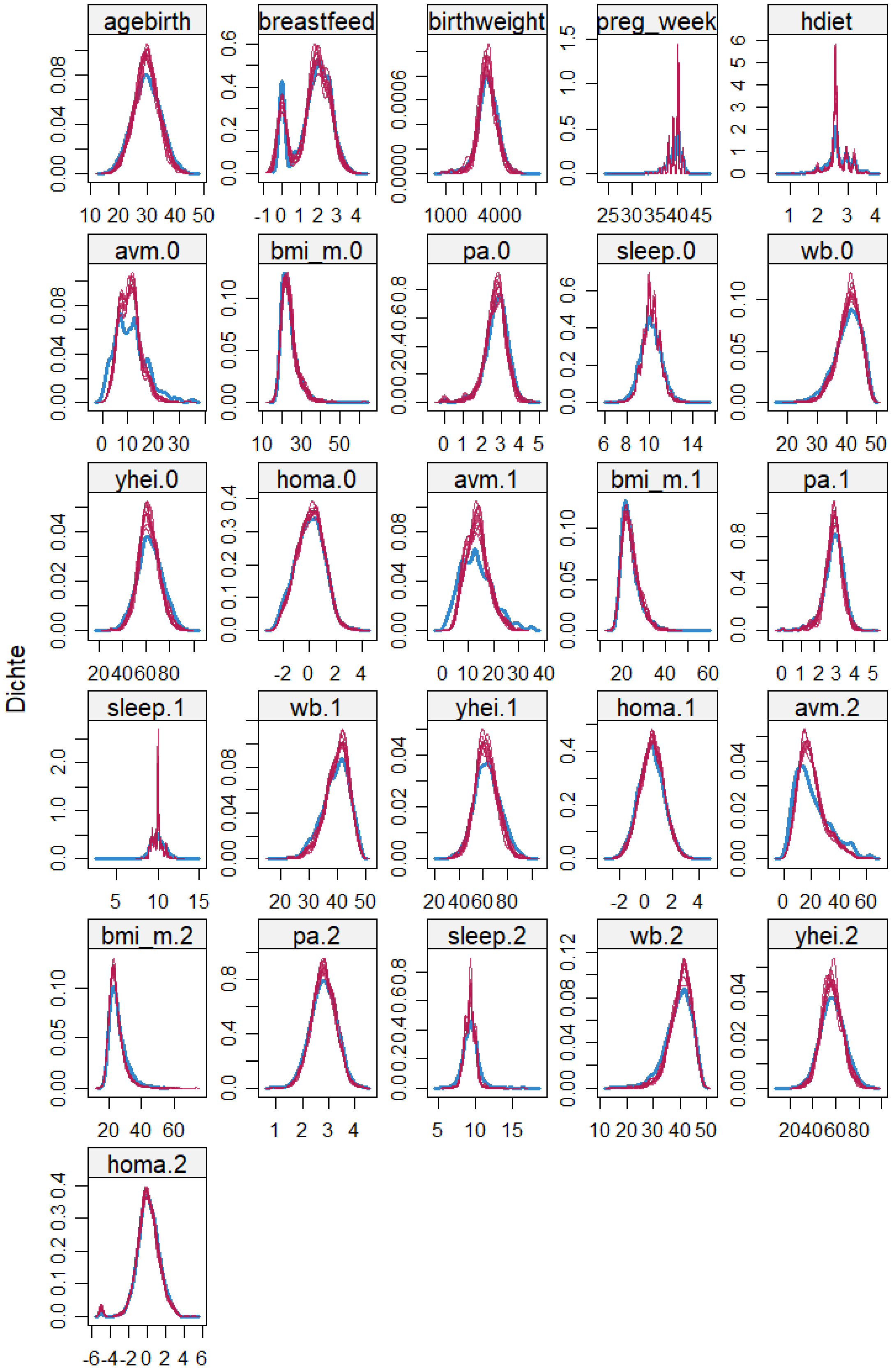
agebirth
breastfeed
birthweight
preg_week
hdiet
avm.0
bmi_m.0
pa.0
sleep.0
wb.0
yhei.0
homa.0
avm.1
bmi_m.1
pa.1
sleep.1
wb.1
yhei.1
homa.1
avm.2
bmi_m.2
pa.2
sleep.2
wb.2
yhei.2
homa.2
Dichte

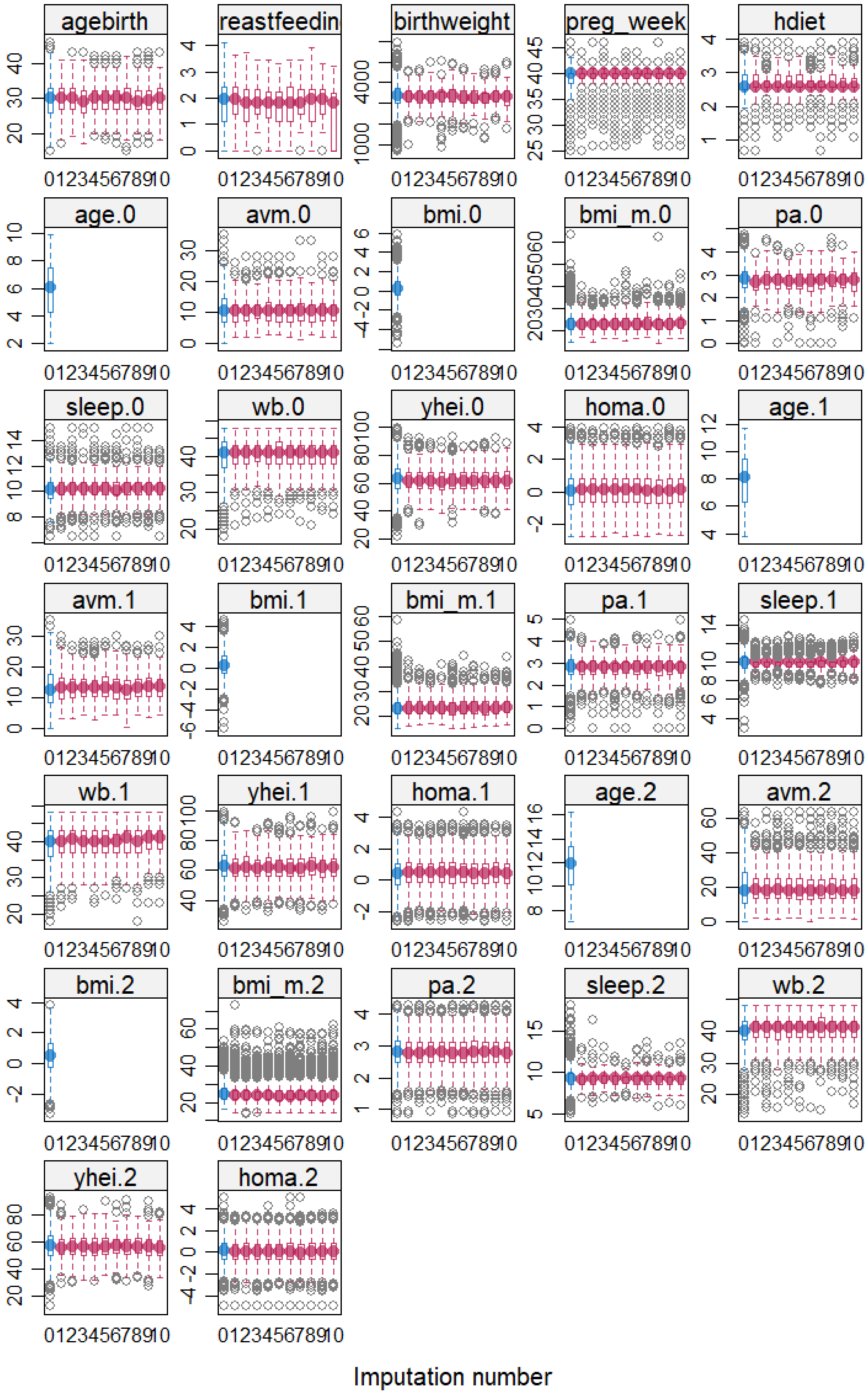
agebirth
reastfeedin
birthweight
preg_week
hdiet
age.0
avm.0
bmi.0
bmi_m.0
pa.0
sleep.0
wb.0
yhei.0
homa.0
age.1
avm.1
bmi.1
bmi_m.1
pa.1
sleep.1
wb.1
yhei.1
homa.1
age.2
avm.2
bmi.2
bmi_m.2
pa.2
sleep.2
wb.2
yhei.2
homa.2
Imputation number

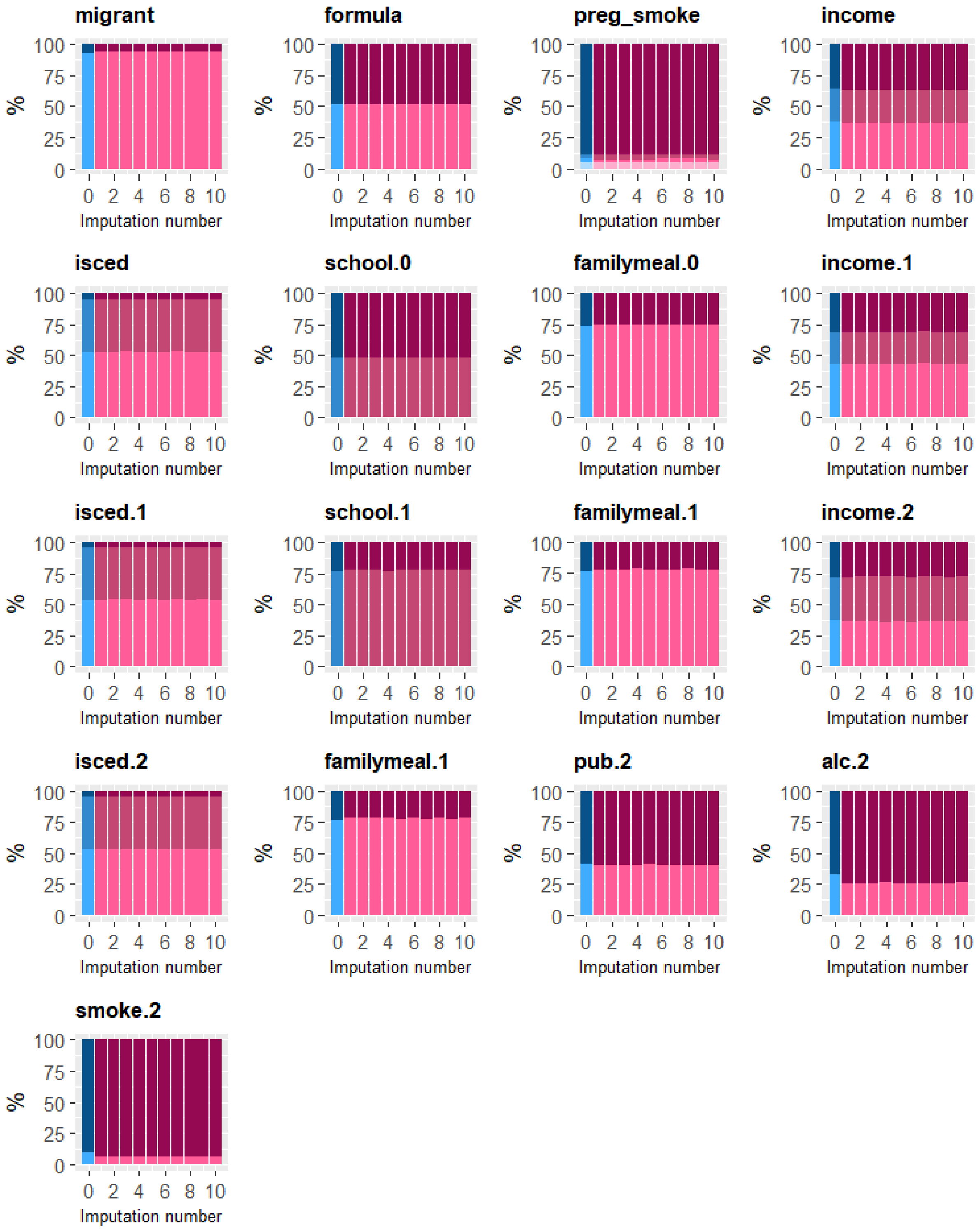


**Figure S2:** Diagnostic plots (kernel density estimates and boxplots for continuous and barplots for discrete variables) of the observed data (blue) and the multiply imputed data (red) with m=10.

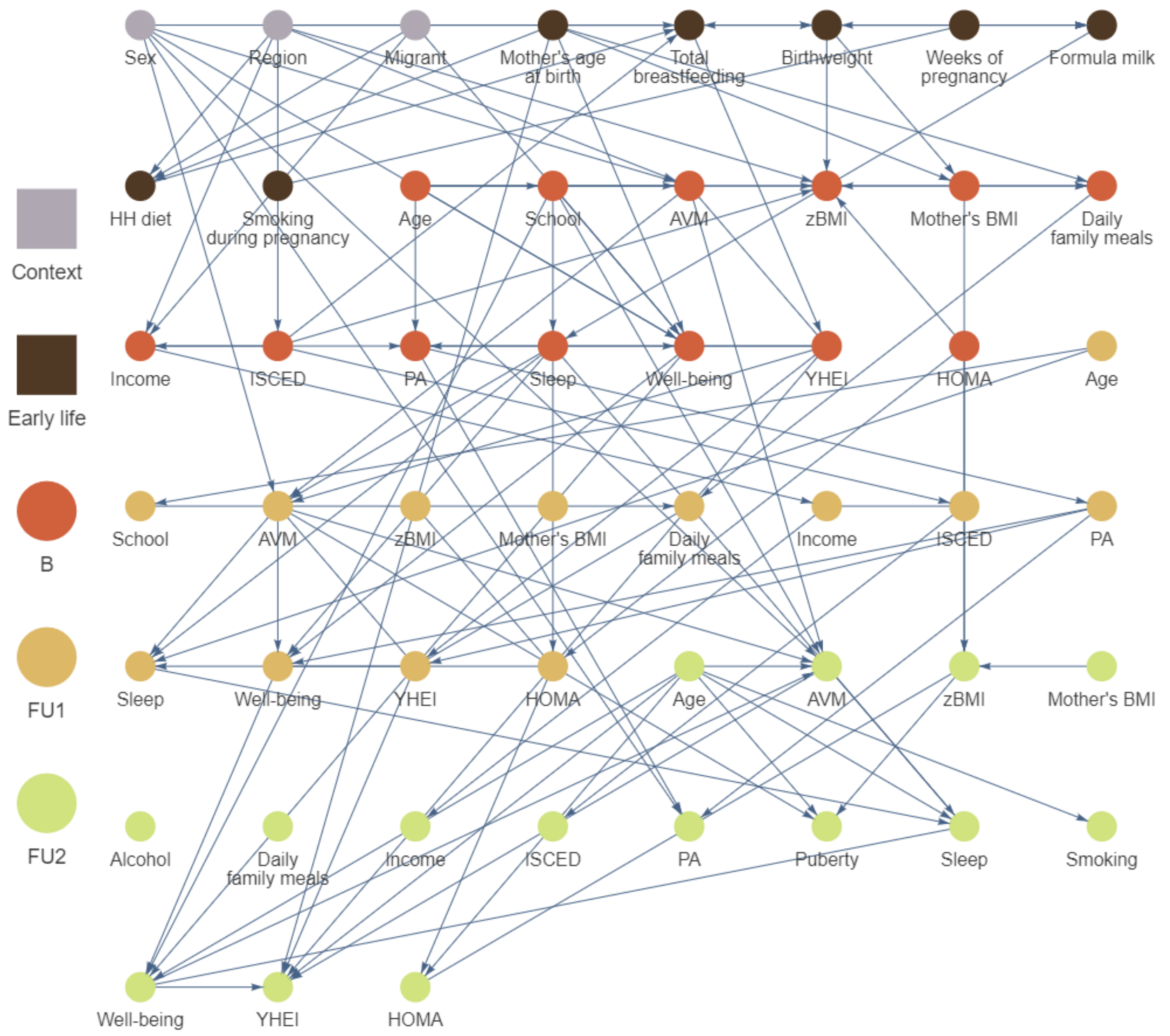


**Figure S3:** Causal graph of childhood obesity based on N = 5,112 European children and adolescents born between 1997 and 2006 estimated by the time-ordered pc-algorithm using **multiple imputation with** $\boldsymbol{\alpha = 0.1}$. Nodes are coloured with respect to their appearance in the life course. Edges without arrowheads could not be orientated by the algorithm.

AVM: audio-visual media consumption, B: Baseline, FU1: first follow-up, FU2: second follow-up, HH diet: month when the child was introduced into the household's diet, HOMA: homeostatic model assessment – insulin resistance, ISCED: highest parental education (International Standard Classification of Education), PA: physical activity, YHEI: youth healthy eating index, zBMI: body mass index z-score

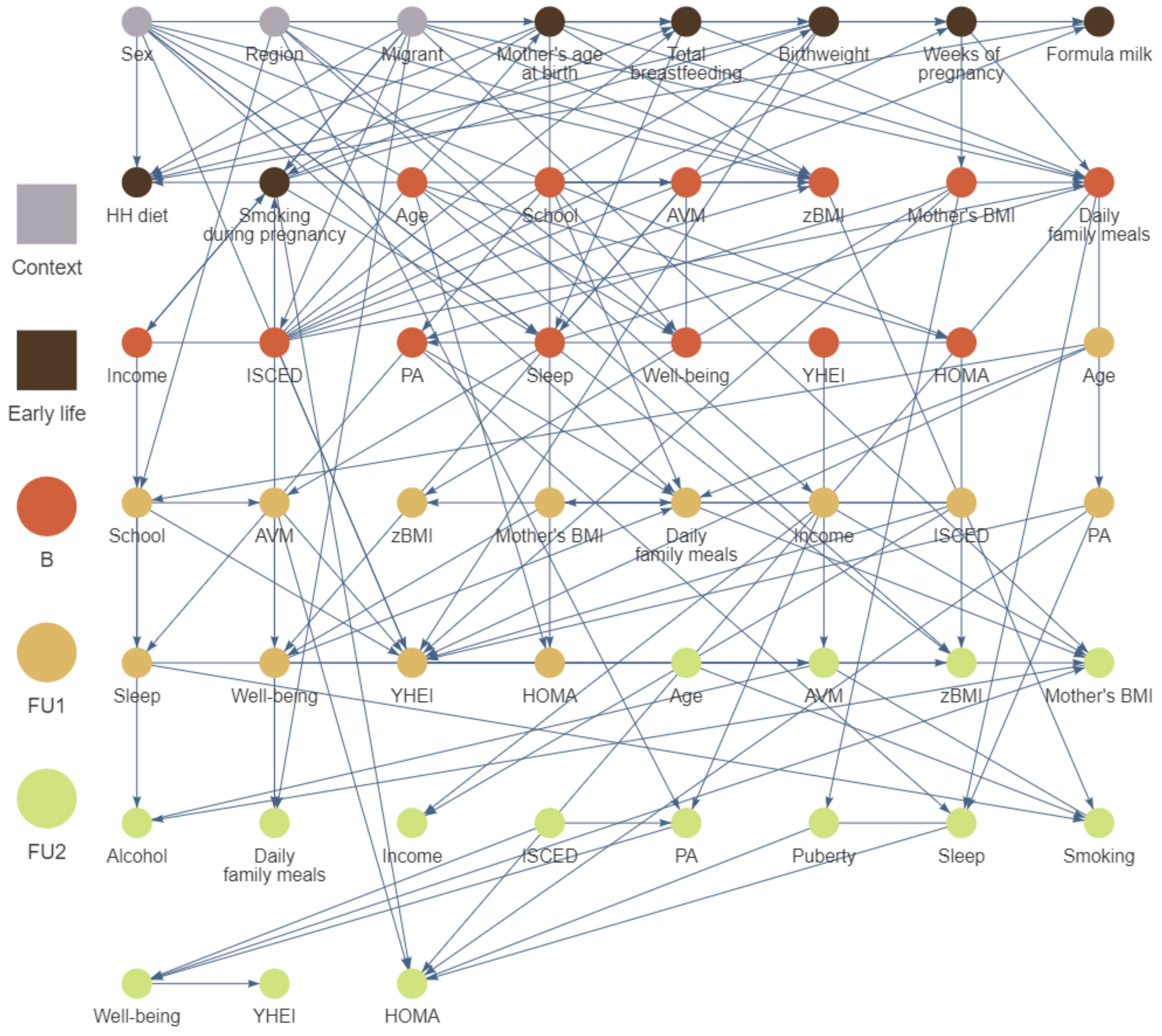


**Figure S4:** Causal graph of childhood obesity based on N = 5,112 European children and adolescents born between 1997 and 2006 estimated by the time-ordered pc-algorithm using **test-wise deletion**. Nodes are coloured with respect to their appearance in the life course. Edges without arrowheads could not be orientated by the algorithm.

AVM: audio-visual media consumption, B: Baseline, FU1: first follow-up, FU2: second follow-up, HH diet: month when the child was introduced into the household's diet, HOMA: homeostatic model assessment – insulin resistance, ISCED: highest parental education (International Standard Classification of Education), PA: physical activity, YHEI: youth healthy eating index, zBMI: body mass index z-score

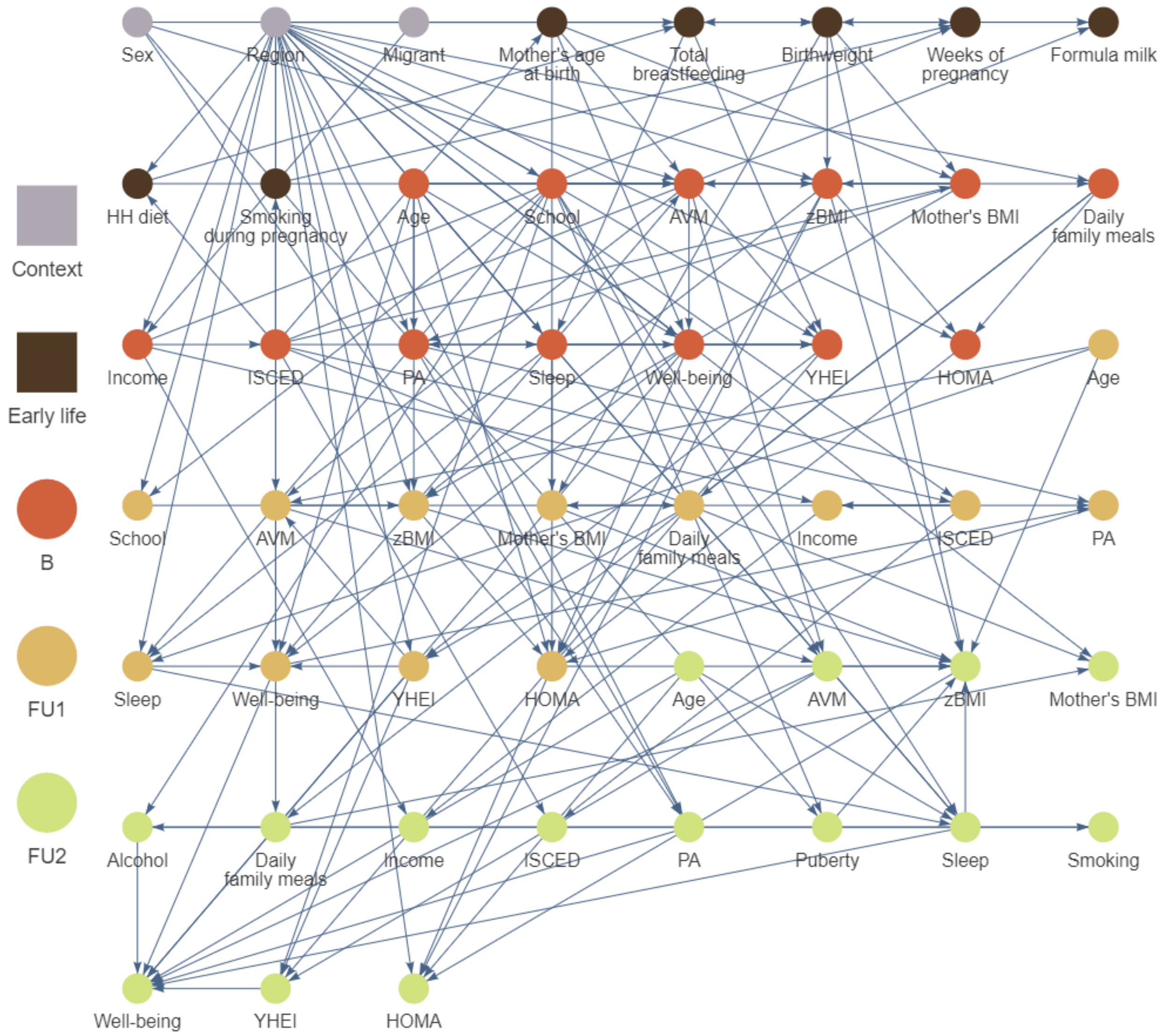


**Figure S5:** Causal graph of childhood obesity based on N = 5,112 European children and adolescents born between 1997 and 2006 estimated by the **Structural EM algorithm**. Nodes are coloured with respect to their appearance in the life course. Edges without arrowheads could not be orientated by the algorithm.

AVM: audio-visual media consumption, B: Baseline, FU1: first follow-up, FU2: second follow-up, HH diet: month when the child was introduced into the household's diet, HOMA: homeostatic model assessment – insulin resistance, ISCED: highest parental education (International Standard Classification of Education), PA: physical activity, YHEI: youth healthy eating index, zBMI: body mass index z-score

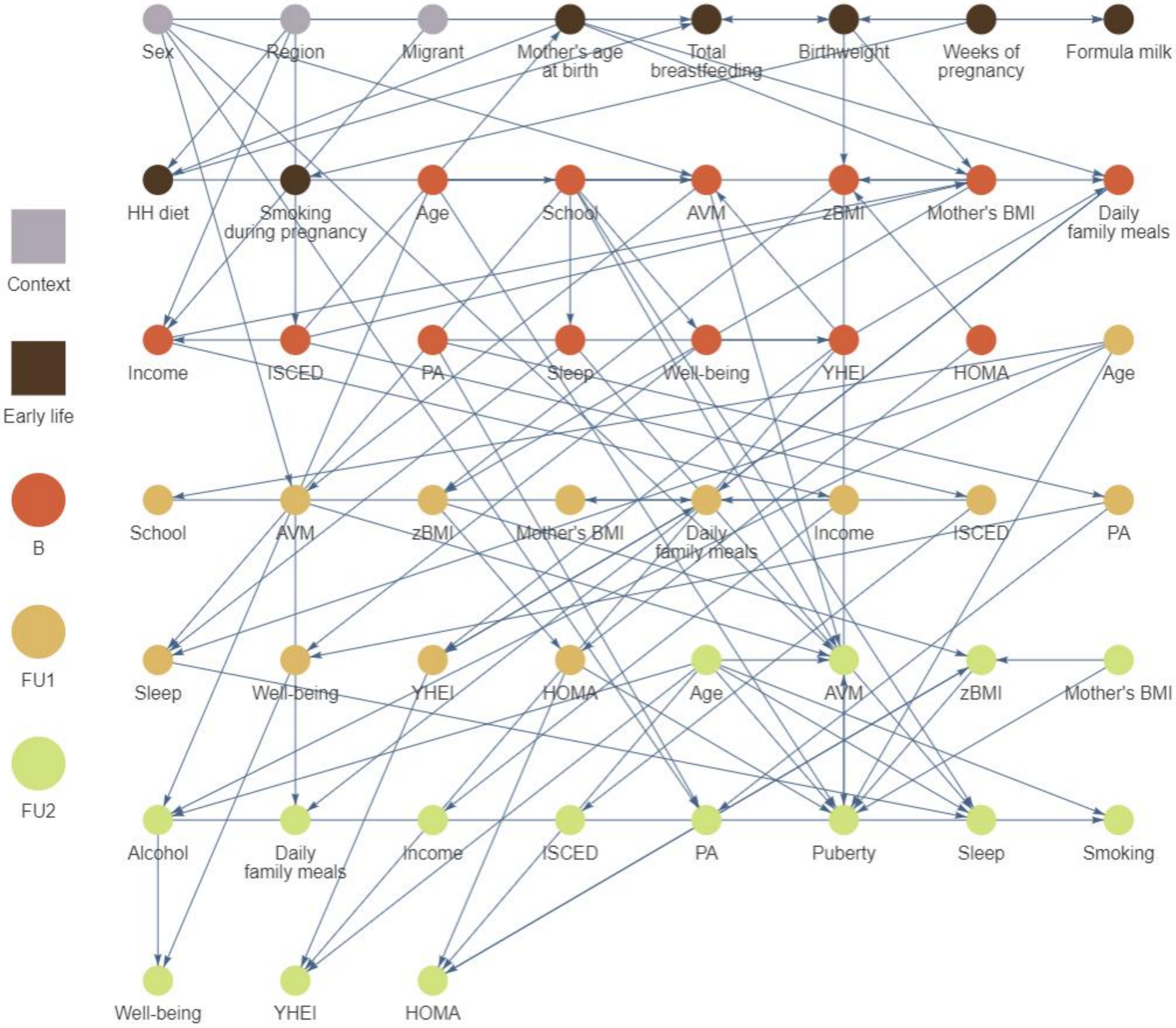


**Figure 6:** Cumulated causal graphs of childhood obesity based on N = 5,112 European children and adolescents born between 1997 and 2006 estimated by the **tiered pc-algorithm** based on one imputed dataset for each of 100 independent **bootstrap** samples. The bootstrap graph contains edges that occurred in more than **44%** of the “bootstrap graphs”. Nodes are coloured with respect to their appearance in the life course. Edges without arrowheads could not be orientated by the algorithm.

AVM: audio-visual media consumption, B: Baseline, FU1: first follow-up, FU2: second follow-up, HH diet: month when the child was introduced into the household's diet, HOMA: homeostatic model assessment – insulin resistance, ISCED: highest parental education (International Standard Classification of Education), PA: physical activity, YHEI: youth healthy eating index, zBMI: body mass index z-score
